# Main COVID-19 information sources in a culturally and linguistically diverse community in Sydney, Australia: A cross-sectional survey

**DOI:** 10.1101/2021.10.24.21265451

**Authors:** J Ayre, DM Muscat, O Mac, C Batcup, E Cvejic, K Pickles, H Dolan, C Bonner, D Mouwad, D Zachariah, U Turalic, Y Santalucia, T Chen, G Vasic, KJ McCaffery

## Abstract

**Background:** Little is known about COVID-19 information-seeking experiences for culturally and linguistically diverse groups in Australia.

**Methods:** Participants were recruited using a cross-sectional survey from March 21 to July 9, 2021, translated into 11 languages, and with supporting bilingual staff. Linear regression models identified factors associated with difficulty finding easy-to-understand COVID-19 information.

**Results:** Across 708 participants (88% born overseas, 31% poor English proficiency), difficulty finding easy-to-understand COVID-19 information was rated 4.13 for English materials (95%CI: 3.85 to 4.41) and 4.36 for translated materials (95%CI: 4.07 to 4.66) (1 easy to 10 hard). Participants who were older (p<0.001), had inadequate health literacy (Mean Difference (MD)=-1.43, 95%CI -2.03 to - 0.82, p<0.001), or poor English proficiency (MD=-1.9, 95%CI-2.51 to -1.29, p<0.001) found it harder to find easy-to-understand English-language COVID-19 information. Those who had greater difficulty finding easy-to-understand translated COVID-19 information were younger (p=0.004), had poor English proficiency (MD=-1.61, 95%CI -2.29 to -0.9, p<0.001), university education (MD=0.77, 95%CI 0.00 to 1.53, p=0.05), and had spent longer living in Australia (p=0.001). They were more likely to rely on friends and family for COVID-19 information (p=0.02). There was significant variation in information-seeking experiences across language groups (p’s<0.001).

**Conclusions:** Easy-to-understand and accessible COVID-19 information is needed to meet the needs of people in culturally and linguistically diverse communities. This approach should involve working alongside these communities to tailor messages and leverage existing communication channels.

## Introduction

Culturally and linguistically diverse communities have endured a disproportionate burden of the COVID-19 pandemic both in Australia and internationally. This is reflected in terms of direct health outcomes (e.g. greater risk of infection and death from COVID-19(Pan et al., 2020)) and psychological and socioeconomic impacts, exacerbated by more crowded living conditions and a larger proportion working in industries not easily adapted to distancing or stay-at-home orders, such as care, healthcare, cleaning, and hospitality (Bambra et al., 2020; van Kooy; Yaya et al., 2020). Adding to this burden, public health communication about COVID-19 has often overlooked the needs of culturally and linguistically communities(Dalzell & Coote, 2021; Viaña et al., 2021). Collaboratively developing tailored, accessible, and understandable communication with these communities is an important step towards equitable healthcare (Wild et al., 2021), but continues to be relatively scarce. Public health communication that is clear and effective across diverse communities is needed to ensure widespread understanding, acceptance, and engagement in COVID-19 prevention behaviours (Habersaat et al., 2020).

The extent that COVID-19 public health communication efforts fall short of community needs can be clearly observed even through relatively crude methods. For example, a recent study showed that the median grade reading score for Australian government COVID-19 information on vaccination, mask-wearing, and distancing ranged from Grade 12 to a university level (Grade 14), for resources collected in April 2021 (Mac et al., 2021) This is 4-6 grades beyond the recommended Grade 8 reading level for effective communication to the average reader in the community(SA Health, 2013). Similarly, our survey of over 4000 Australians at the start of the pandemic (April 2020) found that even single-item questions that roughly estimate health literacy (skills for accessing, understanding and acting on health information) identified that low health literacy was associated with lower confidence understanding government COVID-19 information, and poorer knowledge of COVID-19 symptoms and prevention behaviors [citation removed in anonymized manuscript]. Similar findings were observed for people who did not people speak English as their main language at home [citation removed in anonymized manuscript]. Even when official COVID-19 messages are translated, in Australia these have been criticized for their poor quality(Dalzell S, 2020) and visibility(Grey, 2020).

Several research, service, and policy groups provide guidance on how health organizations can work with communities to create effective communication(Levin-Zamir, 2020; NHS Race and Health Observatory, 2021; White et al., 2021; Wild et al., 2021). For example, Wild and colleagues (2021) emphasize the role of collaboration with the community in tailoring COVID-19 messages for specific communities, and then delivering these messages through trusted messengers using appropriate and accessible channels. Collectively these models advise that public health efforts must disseminate information and advice through communication channels that the community can and do access(Levin-Zamir, 2020; Wild et al., 2021). However, there is limited data to inform how health services identify the most appropriate channels. Our Australia-wide survey conducted in April 2020 found most participants obtained information about COVID-19 through public (Australian government) television (68%), social media (64%), and government websites (64%), but that participants with inadequate health literacy reported that information about COVID-19 was more difficult to find (McCaffery et al., 2020). Though this survey is a useful starting point, participants with inadequate health literacy and who spoke a language other than English at home represented only a small proportion of the total sample; 549 participants (13%) had inadequate health literacy, and only 274 (6%) reported that they did not speak English as their main language at home. The analysis was also limited by the fact that the survey was only available in English, precluding many people from culturally and linguistically diverse communities from taking part. As a result, the study has limited capacity to inform collaborative efforts to tailor COVID-19 public health communication to specific culturally and linguistically diverse communities.

In Australia, areas such as Greater Western Sydney, New South Wales (NSW), are home to dozens of cultural and language groups, with up to 39% of residents born overseas in non-English speaking countries (Public Health Information Development Unit, 2019). This presents real challenges in identifying the most appropriate communication channels as each group may have distinct informational needs and information-seeking behaviors. The current study aimed to explore COVID-19 information-seeking experiences and behaviors across three adjoining regions in Sydney with high cultural and language diversity: Western Sydney, Southwestern Sydney, and Nepean Blue Mountains, between March and July 2021. Data concerning COVID-19 knowledge, attitudes and behaviors were also collected; in this paper we also relate patterns in information-seeking to COVID-19 risk perception, knowledge, and prevention behaviors. Data on COVID-19 testing and vaccination intentions, and the socioeconomic and psychological impacts of the pandemic are reported elsewhere.

## Materials and methods

### Study design

This study used a cross-sectional survey design. The study was approved by Western Sydney Local Health District Human Research Ethics Committee (Project number 2020/ETH03085).

### Setting

Participants were recruited from 21^st^ March to July 9^th^, 2021. During this period, the COVID-19 vaccine rollout had begun across Australia, and daily cases of community transmission in NSW ranged from 0 – 45.(covid19data.com.au, 2021) Restrictions across Greater Sydney began on June 23^rd^ (NSW Health, 2021b), including limitations on the number of people allowed to visit a household, maximum number of people in an exercise class, and reduced seating capacity for outdoor events. On the day the survey closed (July 9^th^) the NSW daily case count was 45, and NSW Health announced stay-at-home orders for Greater Sydney (NSW Health, 2021a). The survey was closed at this time despite some recruitment targets not reached so that results could be more readily interpreted.

### Participants

Participants were eligible to take part if they were aged 18 or over and spoke one of the following as their main language at home: Arabic, Assyrian, Croatian, Dari, Dinka, Hindi, Khmer, Chinese, Samoan, Tongan, or Spanish. We selected these ten language groups through iterative discussions with multicultural health staff, with the aim of providing broad coverage across different global regions, groups with varying average levels of English language proficiency (based on 2016 Australian census data)(Australian Bureau of Statistics, 2016), varying access to translated materials, and varying degrees of reading skill in their main language spoken at home (Appendix 2). Each of the language groups selected was an important group within the Greater Western Sydney region (Western Sydney Local Health District, South Western Sydney Local Health District, Nepean Blue Mountains Local Health District).

Participants were recruited through bilingual Multicultural Health staff and Health Care Interpreter Service staff. Multicultural Health staff recruited participants through their existing networks, community events and community champions. Health Care Interpreter Service staff recruited participants at the end of a medical appointment. Potential participants were offered two means of taking part: Completing the survey themselves online (available in English or translated), or bilingual staff or an interpreter entered responses into the survey platform on the participants’ behalf. To ensure consistency in the phrases used for assisted survey completion, translated versions of the survey were provided to the bilingual staff and interpreters.

### Survey design

Surveys were available in English or translated, by translators with National Accreditation Authority for Translators and Interpreters (NAATI) accreditation where possible. Surveys were hosted on the web-based survey platform Qualtrics. Items relevant to this manuscript are shown in Table 1.

**Table 1.** Survey items.

| Category | Items |
| --- | --- |
| **Socio-demographic** | Self-reported age, gender, education, years living in Australia, main language spoken at home, English language proficiency, reading proficiency in language spoken at home, postcode, access to the internet, access to smartphones, chronic disease, and a single-item health literacy screener<sup>25</sup> |
| **Information sources and information-seeking experiences** | • Top 3 information sources for finding out about COVID-19 in the previous 4 weeks, via 8 categories (TV, radio, social media, websites, printed materials, 'family, friends and community', health professionals, and other. Participants were then asked for more specific answers (e.g. which kind of social media), adapted from our previous COVID-19 survey<sup>14</sup><br>• Which country overseas information came from and the language it was provided in<br>• perceived difficulty finding easy-to-understand information about COVID-19, both in English and in their main language, on a 10-point scale ranging from 1 (not at all difficult) to 10 (extremely difficult) adapted from our previous COVID-19 survey<sup>14</sup> |
| **Knowledge** | Participants asked to identify three signs of COVID-19 and three steps they could take to protect themselves or others from getting COVID-19<sup>14</sup>. Scored out of 6. |
| **Attitudes and intentions** | Risk perception: "how serious a problem do you think COVID-19 is currently, in Australia?" with responses ranging from 0 (not serious at all) to 10 (very serious), adapted from our previous COVID-19 survey<sup>14</sup> |
| **Prevention behaviors** | Participants reflected on the previous four weeks for the following behaviors: I wash my hands frequently with soap and water (for at least 20 seconds); I stay 1.5m away from other people outside my home; I avoid close contact with anyone with cold or flu like symptoms; I have stopped shaking hands; hugging or kissing as a greeting; I wore a mask in places where it was hard to stay 1.5m away from people. captured using 5-point Likert scales (strongly disagree to strongly agree). |

### Analysis plan

Frequencies were weighted (using post-stratification weighting) to reflect each language group’s gender and age group distribution (18-29 years, 30-49 years, 50-69 years, ≥70 years) based on 2016 census data for Western Sydney, South Western Sydney, and Nepean Blue Mountains’ combined populations(Australian Bureau of Statistics, 2016). All summary statistics presented in the results section are weighted unless otherwise indicated. A single participant indicated their gender as ‘other’ and was unable to be included in weighted analyses. Survey items about COVID-19 information sources were re-coded to reflect the categories presented in Table 2, to facilitate a more meaningful interpretation of the results. Multiple linear regression models were used to determine factors associated with risk perception, COVID-19 prevention behaviors (averaged across five behaviors), and knowledge. Age group, gender, health literacy, English-language proficiency, years lived in Australia, risk perception, language group, and information sources were included in each model. The regression also controlled for socioeconomic status of area of residence (based on Index of Relative Socio-economic Advantage and Disadvantage (IRSAD (Australian Bureau of Statistics, 2018)) deciles by postcode), and whether participants completed the survey before or after 23^rd^ June, when restrictions were announced for all of Greater Sydney(NSW Health, 2021b). The IRSAD decile was not available for some participants (n=5), for example, because they had entered digits that did not correspond to a current or previously valid Australian postcode. IRSAD decile for these participants was replaced with the median IRSAD decile for speakers of the same language in the sample. Statistical analysis was conducted using Complex Sample procedures in IBM SPSS Statistics 26.

**Table 2.** Descriptive statistics (categorical variables)

| Variable | n | % |
| --- | --- | --- |
| **Age group** |  |  |
| 18-29 | 147 | 20.8 |
| 30-49 | 295 | 41.7 |
| 50-69 | 193 | 27.3 |
| >70 | 72 | 10.2 |
| **Gender\*** |  |  |
| Male | 344 | 48.7 |
| Female | 363 | 51.3 |
| **Language** |  |  |
| Assyrian | 133 | 18.8 |
| Croatian | 121 | 17.1 |
| Arabic | 80 | 11.3 |
| Chinese | 76 | 10.7 |
| Khmer | 63 | 8.9 |
| Dinka | 63 | 8.9 |
| Dari | 44 | 6.2 |
| Spanish^ | 43 | 6.1 |
| Hindi | 42 | 5.9 |
| Samoan/Tongan | 42 | 5.9 |
| **English language proficiency (How well do you speak English?)** |  |  |
| Very well / well | 487 | 68.9 |
| Not well/not at all | 220 | 31.1 |
| **Literacy in a language other than English (How well do you read in your main language?)** |  |  |
| Very well/ well | 589 | 83.4 |
| Not well/not at all | 118 | 16.6 |
| **Adequate health literacy** | **417** | **58.9** |
| **Highest level of education** |  |  |
| Less than year 12(less than high school) | 115 | 16.2 |
| Year 12 (high school graduate) | 133 | 18.9 |
| Certificate level I to IV / Advanced diploma and diploma level | 249 | 35.3 |
| Bachelor degree level and above | 210 | 29.7 |
| **Has a computer with internet access** | **573** | **81.1** |
| **Has a smartphone** | **686** | **97.1** |
| **Years living in Australia** |  |  |
| 5 years or less | 120 | 16.9 |
| 6 to 10 years | 104 | 14.7 |
| More than 10 years | 398 | 56.4 |
| Born in Australia | 85 | 12.0 |
| **COVID-19 knowledge (correctly naming 3 symptoms and 3 steps to prevent COVID-19 infection for self or others)** | **638** | **90.3** |
| **Total** | **707** |  |

\* 1 respondent indicated 'other/prefer not to say' and is not included in weighted analysis presented in this table; ^ Spanish language group had substantial gaps in recruitment across age groups;

Free-text responses to the question “Are there any cultural practices that you think might mean you are more likely to get COVID-19” were analyzed using content analysis(Weber, 1990). XX familiarized herself with the content and developed a list of preliminary content categories. These categories were refined through discussion with the other authors. XY and XZ coded 305 valid responses according to the final coding framework. The level of agreement was tested with the Cohen Kappa using 40 responses, which indicated substantial agreement (κ=0.83)(Landis & Koch, 1977). Discrepancies were discussed with XW before coding the remaining responses.

## Results

### Sample description

Of the total 708 participants, 442 completed the survey independently (62.4%; unweighted); interpreters completed the survey on behalf of 266 participants (37.6%; unweighted). The mean age was 45.4 years (95% CI: 43.9 to 47.0; range 18–91 years), and 51% of respondents were female (n=363; Table 2). Most participants (88%, n=622) were born in a country other than Australia; 31% reported that they did not speak English well or at all (n=220); 70% had no tertiary qualifications (n=497). Inadequate health literacy was identified for 59% of the sample (n=290). On average, participants rated the difficulty of finding easy-to-understand COVID-19 information in English 4.13 on a scale from 1 (not at all difficult) to 10 (extremely difficult) (95% CI: 3.85 to 4.41). This was on average 4.36 for finding easy-to-understand COVID-19 information in another language (95% CI: 4.07 to 4.66). Almost all participants (90.3%; n=638) could correctly identify 3 symptoms of COVID-19 and 3 steps to prevent the virus’ spread (Table 1). The average score for self-reported adherence to COVID-19 prevention behaviors in the last 4 weeks was 4.40 out of 5 (95% CI:4.32 to 4.47), on a scale from 1 (strongly disagree) to 5 (strongly agree) (Appendix Table S1). When asked to rate how much of a problem COVID-19 is currently in Australia, on average participants responded with 4.37 (where 0 =‘not serious at all’ and 10= ‘very serious’; 95% CI: 4.10 to 4.65) (Appendix Table S2).

### Information sources

The most common information sources for finding out about COVID-19 were official Australian sources/public broadcasters (59.5%, n=421), Australian commercial sources (58.9%, n=417), and social media (56.2%, n=397) (Table 3; Appendix Table S3). TV and websites were the most common formats, with 69.4% of the sample (n=491) reporting that TV was a main information source, and 41.8% reporting using websites (n=296). Overall, half of participants (55.2%, n=390) reported that most of this information was presented in English.

**Table 3.** Main COVID-19 information sources*.

| Main COVID-19 information sources | Total<br>n | % |
| --- | --- | --- |
| **Official Australian source / public broadcaster** | **421** | **59.5** |
| Health professional | 241 | 34.1 |
| Australian public TV | 237 | 33.5 |
| Australian government websites | 163 | 23.1 |
| Australian public radio/podcasts | 57 | 8.1 |
| **Australian Commercial source** | **417** | **58.9** |
| Australian commercial TV | 334 | 47.2 |
| Australian news/magazine website | 140 | 19.8 |
| Australian commercial radio/podcast | 28 | 3.9 |
| Printed newspapers or magazines | 20 | 2.8 |
| **Social media** | **397** | **56.2** |
| Facebook | 312 | 44.1 |
| YouTube | 189 | 26.7 |
| Instagram | 136 | 19.3 |
| WhatsApp | 112 | 15.8 |
| Other (snapchat, twitter, twitch, weibo, wechat) | 124 | 17.6 |
| **Friends or family living in Australia** | **252** | **35.7** |
| Living in Australia more years than participant | 197 | 27.9 |
| Living in Australia same years or fewer | 162 | 22.9 |
| **Community** | **196** | **27.8** |
| Community leader | 99 | 14.0 |
| Community TV | 64 | 9.1 |
| Religious leader | 45 | 6.4 |
| Community radio/podcast | 44 | 6.2 |
| **Overseas information sources** | **205** | **29.0** |
| Overseas website | 128 | 18.2 |
| Overseas TV | 128 | 18.0 |
| Friends or family living overseas | 109 | 15.4 |
| Overseas Radio/podcast | 8 | 1.1 |
| **Any TV** | **491** | **69.4** |
| **Any website** | **296** | **41.8** |

\*Categories and subcategories not mutually exclusive.

The age group with the highest proportion of social media use was for participants aged less than 30 years (78.5%, n=115) (Table 4). Participants in the oldest age group (70 years or more) reported the highest use of friends or family living in Australia (65.6%, n=47), community information sources (including religious or community leaders, community TV and radio) (58.8%, n=42), and overseas information sources (58.6%, n=42). Younger age groups obtained most of their information in English (e.g. 81% of participants <30 years obtained information mostly in English). Participants with inadequate health literacy obtained most of their information in a language other than English (71.0%, n=206); this proportion was 25.6% for participants with adequate health literacy (n=107; Table 4). Participants with inadequate health literacy also reported higher use friends or family (56.8% vs 20.9%), community (38.4% vs 20.9%) and overseas information (41.8% vs 20.2%)

**Table 4.**
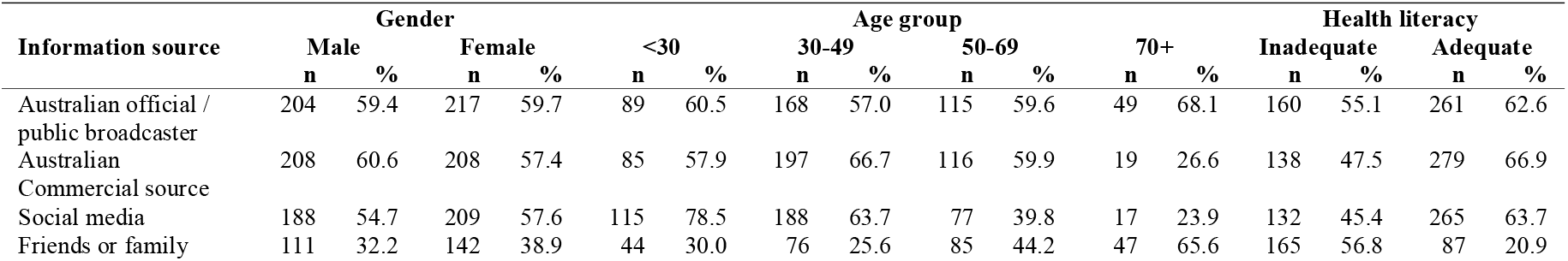
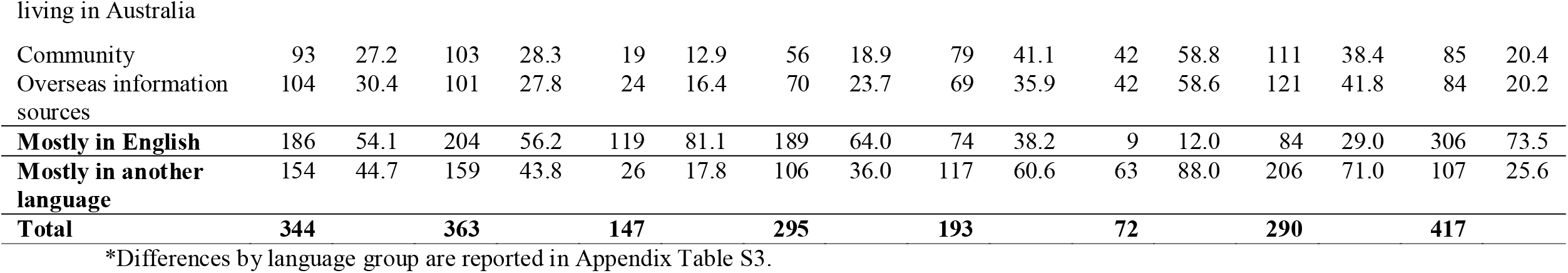
Main COVID-19 information sources, by gender, age group and health literacy*.

| Information source | Gender |  |  |  | Age group |  |  |  |  |  |  |  | Health literacy |  |  |  |
| --- | --- | --- | --- | --- | --- | --- | --- | --- | --- | --- | --- | --- | --- | --- | --- | --- |
|  | Male |  | Female |  | <30 |  | 30-49 |  | 50-69 |  | 70+ |  | Inadequate |  | Adequate |  |
|  | n | % | n | % | n | % | n | % | n | % | n | % | n | % | n | % |
| Australian official / public broadcaster | 204 | 59.4 | 217 | 59.7 | 89 | 60.5 | 168 | 57.0 | 115 | 59.6 | 49 | 68.1 | 160 | 55.1 | 261 | 62.6 |
| Australian Commercial source | 208 | 60.6 | 208 | 57.4 | 85 | 57.9 | 197 | 66.7 | 116 | 59.9 | 19 | 26.6 | 138 | 47.5 | 279 | 66.9 |
| Social media | 188 | 54.7 | 209 | 57.6 | 115 | 78.5 | 188 | 63.7 | 77 | 39.8 | 17 | 23.9 | 132 | 45.4 | 265 | 63.7 |
| Friends or family | 111 | 32.2 | 142 | 38.9 | 44 | 30.0 | 76 | 25.6 | 85 | 44.2 | 47 | 65.6 | 165 | 56.8 | 87 | 20.9 |
| Community | 93 | 27.2 | 103 | 28.3 | 19 | 12.9 | 56 | 18.9 | 79 | 41.1 | 42 | 58.8 | 111 | 38.4 | 85 | 20.4 |
| Overseas information sources | 104 | 30.4 | 101 | 27.8 | 24 | 16.4 | 70 | 23.7 | 69 | 35.9 | 42 | 58.6 | 121 | 41.8 | 84 | 20.2 |
| **Mostly in English** | 186 | 54.1 | 204 | 56.2 | 119 | 81.1 | 189 | 64.0 | 74 | 38.2 | 9 | 12.0 | 84 | 29.0 | 306 | 73.5 |
| **Mostly in another language** | 154 | 44.7 | 159 | 43.8 | 26 | 17.8 | 106 | 36.0 | 117 | 60.6 | 63 | 88.0 | 206 | 71.0 | 107 | 25.6 |
| **Total** | **344** |  | **363** |  | **147** |  | **295** |  | **193** |  | **72** |  | **290** |  | **417** |  |

\*Differences by language group are reported in Appendix Table S3.

Use of Australian official sources or public broadcasters ranged from 28.9% (n=12) for Samoan/Tongan speakers, to 91.7% (n=58) for Khmer speakers (Appendix Table S3). Reliance on family and friends living in Australia as a main source of COVID-19 information ranged from 14.6% (n=9) for Dinka speakers, to 63.3% (n=77) for Croatian speakers; use of community (including TV, radio, and community or religious leaders) ranged from 6.8% (n=5) for Arabic speakers, to 65.3% (n=79) for Croatian speakers. Use of overseas information sources also varied greatly, from 5.3% (n=3) for Khmer speakers, to 98.4% (n=119) for Croatian speakers.

### Analysis: COVID-19 information-seeking experiences, knowledge, risk perception, and prevention behaviors

#### Information-seeking experiences

##### Finding easy-to-understand English-language COVID-19 information

Participants experienced significantly greater difficulty finding easy to understand English-language COVID-19 information if they were older (p<0.001), had inadequate health literacy (Mean Difference (MD)=-1.43, 95%CI -2.03 to -0.82, p<0.001), or had poor English proficiency (MD=-1.9, 95% CI-2.51 to -1.29, p<0.001) (Table 5). There were also differences across language groups (p<0.001; Table 5). Those who reported using Australian commercial information sources reported less difficulty finding easy-to-understand COVID-19 information in English (MD=-0.51, 95% CI -0.94 to -0.08, p=0.02).

**Table 5.** Multiple regression model of factors associated with difficulty finding information about COVID-19 that is easy to understand*.

| Predictor | English-language information |  | Non-English language information |  |
| --- | --- | --- | --- | --- |
|  | Mean difference (95% CI) | P value | Mean difference (95% CI) | P value |
| **Gender** |  |  |  |  |
| Male | Reference |  | Reference |  |
| Female | 0.31 (-0.06 to 0.69) | 0.10 | -0.51 (-1.02 to 0.01) | **0.05** |
| **Age group** |  | **<0.001** |  | **0.004** |
| 18-29 | Reference |  | Reference |  |
| 30-49 | 0.27 (-0.33 to 0.88) | 0.37 | -1.27 (-2.18 to -0.3) | **0.01** |
| 50-69 | 0.29 (-0.43 to 1.01) | 0.43 | -1.71 (-2.72 to -0.6) | **<0.001** |
| >70 | 1.69 (0.74 to 2.64) | **<0.001** | -2.23 (-3.47 to -0.9) | **<0.001** |
| **English-language proficiency** | -1.9 (-2.51 to -1.29) | **<0.001** | -1.61 (-2.29 to -0.9) | **<0.001** |
| **Adequate health literacy** | -1.43 (-2.03 to -0.82) | **<0.001** | 0.13 (-0.53 to 0.79) | 0.70 |
| **Bachelor degree or above education** | -0.17 (-0.71 to 0.37) | 0.54 | 0.77 (0.00 to 1.53) | **0.05** |
| **Risk perception** | 0.03 (-0.05 to 0.11) | 0.42 | -0.08 (-0.19 to 0.03) | 0.15 |
| **Years living in Australia** |  | 0.55 |  | **0.001** |
| 5 years or less | Reference |  | Reference |  |
| 6 to 10 years | 0.13 (-0.52 to 0.79) | 0.69 | 0.93 (0.09 to 1.76) | 0.03 |
| More than 10 years | -0.16 (-0.77 to 0.46) | 0.61 | 1.24 (0.48 to 2.00) | **<0.001** |
| Born in Australia | -0.43 (-1.24 to 0.39) | 0.30 | 1.96 (0.73 to 3.19) | **0.002** |
| **Language<sup>^</sup>** |  | **<0.001** |  | **<0.001** |
| **Information source<sup>†</sup>** |  |  |  |  |
| Official Australian source/public broadcaster | -0.09 (-0.54 to 0.36) | 0.68 | 0.05 (-0.53 to 0.63) | 0.87 |
| Australian commercial source | -0.51 (-0.94 to -0.08) | **0.02** | 0.27 (-0.31 to 0.86) | 0.36 |
| Social media | -0.36 (-0.81 to 0.08) | 0.11 | 0.01 (-0.53 to 0.55) | 0.98 |
| Friends or family living in Australia | -0.23 (-0.69 to 0.23) | 0.33 | 0.70 (0.11 to 1.28) | **0.02** |
| Community | 0.28 (-0.18 to 0.74) | 0.24 | 0.00 (-0.66 to 0.66) | 0.99 |
| Overseas information source | 0.31 (-0.29 to 0.91) | 0.31 | 0.02 (-0.72 to 0.75) | 0.96 |

\*These models also control for IRSAD decile (as a linear variable) and date of survey completion (binary variable, before/after 23 June when restrictions in Greater Sydney were imposed). 1 respondent indicated 'other/prefer not to say' and is not included in weighted analysis;
<sup>^</sup> Individual comparisons for language group not presented; <sup>†</sup> Information sources entered as separate variables as participants could select more than one.

##### Finding easy-to-understand in-language COVID-19 information

Participants who were younger (p=0.004), had poor English proficiency (MD=-1.61, 95% CI -2.29 to -0.9, p<0.001), who had attained a bachelor degree education or higher (MD=0.77, 95% CI 0.00 to 1.53, p=0.05), and who had spent longer living in Australia (p=0.001) experienced significantly greater difficulty finding in-language COVID-19 information that was easy to understand (Table 5). Those who had greater difficulty finding in-language COVID-19 information that was easy to understand were also more likely to rely on friends and family for finding out about COVID-19 (MD=0.70, 95% CI: 0.11 to 1.28, p=0.02). There were also differences observed across language groups (p<0.001; Table 5).

##### Knowledge and risk perception

We observed significant differences in COVID-19 risk perception and knowledge across language groups (p’s<0.001, Table 6). Participants who listed social media as one of their main COVID-19 information sources reported significantly higher knowledge (MD=0.29, 95% CI: 0.05 to 0.54, p=0.02) and risk perception (MD =0.56, 95% CI: 0.11 to 1.02, p=0.02), compared to those who did not use social media. English-language proficiency, health literacy, and education were not significantly associated with COVID-19 knowledge or risk perception. Female participants obtained lower knowledge scores compared to males (OR=-0.20, 95%CI: -0.40 to 0.00, p=0.05). Participants who had lived in Australia for more than 10 years had lower risk perception compared to participants who had lived in Australia 5 years or less (OR=0.82, 95%CI: 0.22 to 1.42, p=0.01).

**Table 6.** Multiple linear regression model of factors associated with knowledge, risk perception and behaviors*.

| Predictor | Knowledge (out of 6) |  | Risk perception |  | Behaviors |  |
| --- | --- | --- | --- | --- | --- | --- |
|  | Mean difference (95% CI) | P value | Mean difference (95% CI) | P value | Mean difference (95% CI) | P value |
| **Gender** |  |  |  |  |  |  |
| Male | Reference |  | Reference |  |  | Reference |
| Female | -0.20 (-0.40 to 0.0) | **0.05** | 0.27 (-0.14 to 0.67) | 0.20 | 0.04 (-0.08 to 0.17) | 0.49 |
| **Age group** |  | 0.53 |  | **0.05** |  | **<0.001** |
| 18-29 | Reference |  | Reference |  | Reference |  |
| 30-49 | 0.11 (-0.18 to 0.40) | 0.45 | -0.63 (-1.28 to 0.02) | 0.06 | 0.07 (-0.11 to 0.24) | 0.44 |
| 50-69 | 0.14 (-0.23 to 0.51) | 0.46 | -0.77 (-1.56 to 0.02) | 0.06 | 0.32 (0.13 to 0.51) | **<0.001** |
| >70 | -0.08 (-0.56 to 0.41) | 0.76 | -0.17 (-1.10 to 0.76) | 0.72 | 0.44 (0.21 to 0.68) | **<0.001** |
| **English-language proficiency** | 0.06 (-0.21 to 0.33) | 0.67 | -0.19 (-0.77 to 0.40) | 0.53 | 0.07 (-0.07 to 0.22) | 0.32 |
| **Adequate health literacy** | 0.06 (-0.19 to 0.31) | 0.65 | -0.06 (-0.65 to 0.54) | 0.86 | 0.10 (-0.06 to 0.26) | 0.22 |
| **Bachelor degree or above education** | -0.02 (-0.32 to 0.28) | 0.91 | 0.08 (-0.47 to 0.63) | 0.79 | -0.04 (-0.25 to 0.16) | 0.68 |
| **Risk perception** | 0.05 (0.00 to 0.10) | **0.04** | — |  | 0.05 (0.02 to 0.07) | **<0.001** |
| **Years living in Australia** |  | 0.16 |  | **0.001** |  | **<0.001** |
| 5 years or less | Reference |  | Reference |  | Reference |  |
| 6 to 10 years | -0.04 (-0.63 to 0.55) | 0.90 | 0.19 (-0.50 to 0.88) | 0.59 | -0.30 (-0.52 to -0.00) | **0.01** |
| More than 10 years | 0.10 (-0.41 to 0.62) | 0.69 | 0.82 (0.22 to 1.42) | **0.01** | -0.40 (-0.57 to -0.20) | **<0.001** |
| Born in Australia | -0.02 (-0.74 to 0.70) | 0.96 | -0.54 (-1.36 to 0.28) | 0.20 | -0.45 (-0.71 to -0.20) | **<0.001** |
| **Language<sup>^</sup>** |  | **<0.001** |  | **<0.001** |  | **<0.001** |
| **Information source<sup>†</sup>** |  |  |  |  |  |  |
| Official Australian source/public broadcaster | 0.24 (0.00 to 0.48) | **0.05** | 0.09 (-0.42 to 0.60) | 0.72 | 0.01 (-0.14 to 0.16) | 0.91 |
| Australian commercial source | -0.11 (-0.34 to 0.12) | 0.36 | 0.19 (-0.27 to 0.66) | 0.41 | 0.15 (-0.04 to 0.34) | 0.11 |
| Social media | 0.29 (0.05 to 0.54) | **0.02** | 0.56 (0.11 to 1.02) | **0.02** | 0.02 (-0.12 to 0.16) | 0.74 |
| Friends or family living in Australia | 0.16 (-0.06 to 0.37) | 0.15 | 0.68 (0.17 to 1.19) | **0.01** | -0.13 (-0.25 to -0.00) | **0.04** |
| Community | 0.07 (-0.20 to 0.33) | 0.62 | 0.30 (-0.30 to 0.90) | 0.33 | 0.11 (-0.06 to 0.29) | 0.21 |
| Overseas information source | 0.01 (-0.28 to 0.3) | 0.95 | -0.28 (-1.02 to 0.47) | 0.47 | -0.10 (-0.29 to 0.08) | 0.28 |

\*These models also control for IRSAD decile (as a linear variable) and date of survey completion (binary variable, before/after 23 June when restrictions in Greater Sydney were imposed). 1 respondent indicated 'other/prefer not to say' and is not included in weighted analysis; <sup>^</sup> Individual comparisons for language group not presented; <sup>†</sup> Information sources entered as separate variables as participants could select more than one.

##### COVID-19 prevention behaviors

English-language proficiency, health literacy, and education were not significantly associated with adherence to COVID-19 prevention behaviors (Table 6). Years lived in Australia was inversely associated with COVID-19 prevention behaviors (p<0.001); participants who had lived in Australia for 6 to 10 years, more than 10 years, or who were born in Australia, reported lower levels of COVID-19 prevention behaviors compared to participants who had moved to Australia in the previous 5 years. We observed differences in self-reported COVID-19 prevention behaviors across language groups (p<0.001). Relying on family and friends living in Australia as a source of information for COVID-19 was associated with significantly lower self-reported COVID-19 prevention behaviors (MD =-0.13, 95% CI: -0.25 to -0.00, p=0.04).

Participants were asked to describe what cultural practices might increase the spread of COVID-19. From the 305 responses (43.2%) we generated 6 topics to categories responses (Table 7). For example, 61.4% of the participants who provided a response (n=188) indicated that greetings made distancing behaviors more challenging, followed by community gatherings (43.8%, n=134).

**Table 7.** Content analysis of practices perceived to increase COVID-19 infection (n=305)*

| **Topic** | **Example quote** | **n** | **%** |
| --- | --- | --- | --- |
| Greetings | *"The way we greet each other, lots of contact and physical closeness"* | 188 | 61.4 |
| Gatherings | *"Our soccer games, church gatherings, and the way we greet - hand shaking, kissing and hugging"* | 134 | 43.8 |
| Attending a place of worship | *"Participation in weddings and religious ceremonies"* | 30 | 9.8 |
| Close proximity to others | *"Stay close to the sick people and no mask"* | 26 | 8.6 |
| Not wearing masks | *"Many people refuse to wear masks, and they are unfriendly towards people who do wear masks."* | 8 | 2.8 |
| Social home visit | *"visiting relative and family gathering, and people are hiding any symptoms"* | 6 | 1.9 |

\*Responses could be coded to more than 1 Topic

## Discussion

In this Australian survey of people from culturally and linguistically diverse communities, participants who were older, who had inadequate health literacy and poor self-reported English proficiency found it harder to find English-language COVID-19 information that was easy for them to understand. Those who had greater difficulty finding easy-to-understand translated COVID-19 information were more likely to rely on friends and family for this information. Using social media to find out about COVID-19 was more common for younger participants, whereas more than half of participants aged over 70 years relied on friends or family living in Australia (66%), community (59%), and overseas information sources (59%). More than half of participants with inadequate health literacy used friends or family living in Australia as a main information source (57%). Perceived seriousness of COVID-19 (risk perception), and difficulty finding easy-to-understand information about COVID-19 differed across language groups. Most participants could correctly identify three COVID-19 symptoms and three steps to prevent its spread, and self-reported high adherence to COVID-19 prevention behaviors. Use of social media as a main COVID-19 information source was associated with greater knowledge and risk perception; more years living in Australian and relying on Australian friends and family for information about COVID-19 were both associated with lower prevention behaviors.

The high level of variation across language groups emphasizes the need for tailored public communication efforts that meet the needs of people in the community, not only in terms of translation, but also health literacy and consideration of communication channel accessibility and preferences(Wild et al., 2021). Some communication channels may be underutilized. For example, less than one third of participants listed community resources (including leaders, TV and radio) as a main information source, despite the clear role that these resources play in effective communication and increased engagement (NHS Race and Health Observatory, 2021; Wild et al., 2021). They were also more often used by groups who had difficulty finding information that was easy to understand (older age and inadequate health literacy). Increased funding, resources, policy and infrastructure are needed to establish, support and strengthen community-based communication channels. Similarly, only one-third of participants reported health professionals as a main information source. Actively engaging health professionals and community health to deliver public health messages may be another avenue for improving COVID-19 communication, as suggested by the UK’s National Health Service Race and Health Observatory COVID-19 Working Group (NHS Race and Health Observatory, 2021).

There is also opportunity to augment COVID-19 communication channels that are already widely used, for example, social media. Whilst social media is often discussed as a contributor to misinformation (Islam et al., 2020), in this study, it was associated with greater COVID-19 knowledge and perceived seriousness of COVID-19. Similarly, US research suggests that social media may be a useful COVID-19 communication channel for Spanish-speaking Americans (Ramos et al., 2020). Public and commercial TV were also widely used in our sample but could be made more inclusive by consistently incorporating subtitles. Efforts could also ensure that information shared amongst friends and family is of high quality and accuracy, as we found this information source was associated with lower self-reported COVID-19 prevention behaviors.

Two findings from this survey warrant further comment. Firstly, that people who were more educated reported it harder to find easy-to-understand translated COVID-19 information. This could reflect a desire for more detailed or complex Australian COVID-19 information (which is often not translated into other languages), or that this group has less contact with organizations that disseminate more detailed translated information (e.g. community organizations). Secondly, participants who had lived in Australia for fewer years reported greater adherence to COVID-19 prevention behaviors. One possible interpretation is that more recent migrants are more concerned about fines or legal notices for violating public health orders because of the perceived implications for visa security.

### Strengths and limitations

This study is strengthened by recruitment methods that are inclusive and reduce barriers to participation, such as translated versions of the survey, use of interpreters, and use of multiple recruitment methods (including through social media, community events, and through community networks). Further, by including several variables related to culture and language (e.g. English language proficiency, literacy in own language, and years living in Australia), and focusing on 10 specific language groups (more detail provided in our community summaries) (Appendix 3), this study provides a more nuanced understanding of the sample, providing more practical avenues of action to support these communities. This is in stark contrast to many studies which are only able to provide data on e.g. language spoke at home or years living in Australia, including our own previous work (McCaffery et al., 2020). Further work could explore experiences within a single language or culture to provide even more specific practical avenues of action.

The limitations of the study are that recruitment for some language groups was lower than anticipated (n<50). For these language groups, estimates may be less reliable. In addition, using a relatively simple knowledge measure, we observed high levels of knowledge and self-reported COVID-19 prevention behaviors, and this may have limited our ability to identify important predictors of these outcomes. Future work could consider more difficult knowledge questions or objectively observe behaviors. Practical constraints also limited the number of languages we could include; further research involving other languages and cultures in Greater Western Sydney (and beyond) will deepen our understanding of how people in these communities have experienced the COVID-19 pandemic in Australia.

### Conclusion

Culturally and linguistically diverse communities in Greater Western Sydney each have distinct patterns of COVID-19 risk, behaviors, and information-seeking experiences and preferences. This study highlights that across languages, those who speak English less proficiently and who have inadequate health literacy find it more challenging to find information about COVID-19 that is easy to understand. Community and health professional communication channels could benefit from additional support that increases their capacity to engage and inform people in these communities. Efforts could also focus on ensuring the quality and accuracy of information spread by worth of mouth through family and friends.

## Data Availability

Data available upon reasonable request to the authors

## Acknowledgements

We would like to acknowledge the efforts of all community health workers, local health district staff, community champions, and community leaders who supported and contributed to this project. We would also like to thank all participants for their involvement.

## Data availability statement

Data available upon request from author

