## Appendix Table for "Main COVID-19 information sources in a culturally and linguistically diverse community in Sydney, Australia: A cross-sectional survey"

### Appendix 1: Supplementary tables

**Table S1. Descriptive characteristics, difficulty finding information and risk perception***

|  | **Difficulty finding COVID-19 information in English that is easy to understand  ( 1 low to 10 high)** | | **Difficulty finding COVID-19 information in a language other than English that is easy to understand  ( 1 low to 10 high)** | | **Knowledge of COVID-19 symptoms and steps to stop the virus’ spread (0 low to 6 high)** | |
| --- | --- | --- | --- | --- | --- | --- |
| **Variable** | **M** | **SE** | **M** | **SE** | **M** | **SE** |
| **Age group** |  |  |  |  |  |  |
| 18-29 | 2.93 | 0.34 | 5.46 | 0.47 | 5.63 | 0.12 |
| 30-49 | 3.44 | 0.19 | 4.20 | 0.24 | 5.61 | 0.08 |
| 50-69 | 4.80 | 0.23 | 4.03 | 0.21 | 5.60 | 0.09 |
| >70 | 7.59 | 0.31 | 3.68 | 0.28 | 5.54 | 0.14 |
| **Gender** |  |  |  |  |  |  |
| Male | 3.96 | 0.21 | 4.64 | 0.24 | 5.69 | 0.07 |
| Female | 4.29 | 0.19 | 4.10 | 0.18 | 5.53 | 0.07 |
| **Language** |  |  |  |  |  |  |
| Arabic | 3.82 | 0.44 | 2.81 | 0.59 | 5.83 | 0.09 |
| Assyrian | 4.15 | 0.32 | 5.57 | 0.34 | 5.18 | 0.16 |
| Chinese | 4.09 | 0.45 | 3.35 | 0.38 | 5.64 | 0.13 |
| Croatian | 6.55 | 0.24 | 3.36 | 0.19 | 6.00 | 0.00 |
| Dari | 7.04 | 0.49 | 6.99 | 0.54 | 5.87 | 0.13 |
| Dinka | 2.60 | 0.34 | 5.62 | 0.49 | 5.05 | 0.27 |
| Hindi | 1.72 | 0.24 | 3.25 | 0.53 | 5.97 | 0.03 |
| Khmer | 3.12 | 0.37 | 2.79 | 0.26 | 5.78 | 0.13 |
| Spanish^^^ | 2.89 | 0.37 | 5.21 | 0.70 | 5.51 | 0.19 |
| Samoan/Tongan | 2.19 | 0.33 | 6.18 | 0.50 | 5.39 | 0.28 |
| **English language proficiency (How well do you speak English?)** | | | |  |  |  |
| Very well/ well | 2.88 | 0.14 | 4.34 | 0.20 | 5.58 | 0.07 |
| Not well/not at all | 6.90 | 0.20 | 4.40 | 0.22 | 5.67 | 0.07 |
| **Literacy in a language other than English (How well do you read in your main language?)** | | | |  |  |  |
| Very well/ well | 4.24 | 0.15 | 4.04 | 0.15 | 5.63 | 0.05 |
| Not well/not at all | 3.58 | 0.36 | 5.96 | 0.45 | 5.51 | 0.15 |
| **Health literacy** |  |  |  |  |  |  |
| Inadequate | 6.22 | 0.24 | 4.27 | 0.20 | 5.61 | 0.07 |
| Adequate | 2.68 | 0.14 | 4.42 | 0.22 | 5.60 | 0.07 |
| **Years living in Australia** |  |  |  |  |  |  |
| 5 years or less | 4.63 | 0.37 | 3.86 | 0.38 | 5.72 | 0.10 |
| 6 to 10 years | 4.32 | 0.34 | 4.05 | 0.36 | 5.69 | 0.10 |
| More than 10 years | 4.31 | 0.18 | 4.34 | 0.19 | 5.53 | 0.07 |
| Born in Australia | 2.38 | 0.33 | 5.54 | 0.60 | 5.71 | 0.14 |
| **Total** | **4.13** | **0.14** | **4.36** | **0.15** | **5.61** | **0.05** |

* 1 respondent indicated ‘other/prefer not to say’ and is not included in weighted analysis presented in this table; ^ Spanish language group had substantial gaps in recruitment across age groups;

**Table S2. Descriptive characteristics, risk perception and COVID-19 prevention behaviours***

|  | **COVID-19 risk perception in Australia (0 low to 10 high)** | | **COVID-19 prevention behaviour**  **(1 low to 5 high)** | |
| --- | --- | --- | --- | --- |
| **Variable** | **M** | **SE** | **M** | **SE** |
| **Age group** |  |  |  |  |
| 18-29 | 4.56 | 0.41 | 4.36 | 0.09 |
| 30-49 | 4.34 | 0.22 | 4.35 | 0.07 |
| 50-69 | 4.06 | 0.19 | 4.48 | 0.05 |
| >70 | 4.95 | 0.30 | 4.44 | 0.06 |
| **Gender** |  |  |  |  |
| Male | 4.16 | 0.22 | 4.38 | 0.05 |
| Female | 4.57 | 0.17 | 4.42 | 0.06 |
| **Language** |  |  |  |  |
| Arabic | 2.27 | 0.31 | 4.72 | 0.08 |
| Assyrian | 3.15 | 0.23 | 4.28 | 0.08 |
| Chinese | 2.80 | 0.29 | 4.70 | 0.06 |
| Croatian | 4.00 | 0.18 | 4.11 | 0.05 |
| Dari | 7.92 | 0.54 | 4.91 | 0.03 |
| Dinka | 5.51 | 0.46 | 4.24 | 0.16 |
| Hindi | 6.32 | 0.67 | 4.21 | 0.22 |
| Khmer | 7.31 | 0.22 | 4.86 | 0.05 |
| Spanish^^^ | 4.75 | 0.74 | 3.80 | 0.30 |
| Samoan/Tongan | 4.00 | 0.51 | 4.28 | 0.12 |
| **English language proficiency (How well do you speak English?)** | | | |  |
| Very well/ well | 4.30 | 0.18 | 4.37 | 0.05 |
| Not well/not at all | 4.53 | 0.21 | 4.47 | 0.04 |
| **Literacy in a language other than English (How well do you read in your main language?)** | | | |  |
| Very well/ well | 4.32 | 0.15 | 4.42 | 0.04 |
| Not well/not at all | 4.61 | 0.37 | 4.30 | 0.10 |
| **Health literacy** |  |  |  |  |
| Inadequate | 4.50 | 0.20 | 4.41 | 0.04 |
| Adequate | 4.28 | 0.19 | 4.39 | 0.06 |
| **Years living in Australia** |  |  |  |  |
| 5 years or less | 3.70 | 0.39 | 4.69 | 0.05 |
| 6 to 10 years | 4.73 | 0.36 | 4.49 | 0.15 |
| More than 10 years | 4.71 | 0.17 | 4.34 | 0.05 |
| Born in Australia | 3.30 | 0.43 | 4.14 | 0.10 |
| **Total** | **4.37** | **0.14** | **4.40** | **0.38** |

* 1 respondent indicated ‘other/prefer not to say’ and is not included in weighted analysis presented in this table; ^ Spanish language group had substantial gaps in recruitment across age groups;

**Table S3. Main COVID-19 information sources, by language group***

| **Information Source** | **Arabic** | | **Assyrian** | | **Croatian** | | **Dari** | | **Dinka** | | **Hindi** | | **Khmer** | | **Chinese** | | **Samoan/ Tongan** | | **Spanish^^^** | |
| --- | --- | --- | --- | --- | --- | --- | --- | --- | --- | --- | --- | --- | --- | --- | --- | --- | --- | --- | --- | --- |
|  | **n** | **%** | **n** | **%** | **n** | **%** | **n** | **%** | **n** | **%** | **n** | **%** | **n** | **%** | **n** | **%** | **n** | **%** | **n** | **%** |
| Official Australian source / public broadcaster | 39 | 48.6 | 77 | 57.8 | 73 | 60.4 | 30 | 67.2 | 42 | 66.5 | 29 | 68.5 | 58 | 91.7 | 34 | 44.2 | 12 | 28.9 | 28 | 65.7 |
| Australian Commercial source | 39 | 48.6 | 84 | 63.3 | 56 | 46.6 | 17 | 38.7 | 35 | 54.8 | 31 | 73.7 | 52 | 81.8 | 35 | 46.1 | 34 | 81.9 | 33 | 77.8 |
| Social media | 54 | 68.1 | 55 | 41.0 | 46 | 38.3 | 32 | 73.2 | 41 | 65.2 | 30 | 71.1 | 36 | 56.6 | 54 | 71.5 | 21 | 49.0 | 28 | 65.2 |
| Friends or family living in Australia | 15 | 18.2 | 43 | 32.4 | 77 | 63.3 | 24 | 53.7 | 9 | 14.6 | 6 | 15.2 | 21 | 32.6 | 35 | 46.7 | 7 | 16.7 | 16 | 36.1 |
| Community | 5 | 6.8 | 30 | 22.8 | 79 | 65.3 | 3 | 7.7 | 16 | 25.1 | 9 | 21.9 | 22 | 35.4 | 14 | 18.7 | 12 | 27.5 | 5 | 11.9 |
| Overseas information sources | 10 | 12.6 | 11 | 8.6 | 119 | 98.4 | 4 | 8.7 | 3 | 4.9 | 14 | 32.8 | 3 | 5.3 | 25 | 33.1 | 3 | 6.6 | 4 | 10.1 |
| **Mostly in English** | 51 | 63.7 | 85 | 63.8 | 2 | 1.8 | 19 | 42.4 | 60 | 94.6 | 36 | 84.9 | 38 | 60.2 | 32 | 41.5 | 34 | 81.0 | 35 | 81.4 |
| **Mostly in another language** | 29 | 36.3 | 44 | 33.3 | 119 | 98.2 | 25 | 57.6 | 3 | 5.4 | 6 | 15.1 | 25 | 39.8 | 44 | 58.5 | 8 | 19.0 | 8 | 18.6 |
| **Total** | **80** |  | **133** |  | **121** |  | **44** |  | **63** |  | **42** |  | **63** |  | **76** |  | **42** |  | **43** |  |

* 1 respondent indicated ‘other/prefer not to say’ and is not included in weighted analysis presented in this table; ^ Spanish language group had substantial gaps in recruitment across age groups;
