## Appendix 2 for "Main COVID-19 information sources in a culturally and linguistically diverse community in Sydney, Australia: A cross-sectional survey"

### Appendix 2: Language selection

|  | | **Greater Western Sydney (2016) census data** | **NSW (2016) census data** | | | | | |
| --- | --- | --- | --- | --- | --- | --- | --- | --- |
|  | **Language spoken at home** | **Population** | **% low English proficiency** | **% born in Australia** | **% born overseas who arrived in last 5 years (2011 to 2016)** | **Availability of translated health materials in NSW*** | **% with tertiary qual (university)** | **% in low income households (earning <$600/week)** |
| **Category 1:**   - Adequate literacy in their own language* - High English proficiency (≤10% low) | Hindi | 47,924 | 4.5 | 15.5 | 28.3 | Common | 52.7 | 10.3 |
|  | Pacific Islander (Samoan/ Tongan) | Samoan: 14,506  Tongan: 7,441 | Samoan: 8.3  Tongan: 9.3 | Samoan: 20.4  Tongan: 34.7 | Samoan: 24.2  Tongan: 16.2 | Uncommon | Samoan: 4.3  Tongan: 6.0 | Samoan: 14.6  Tongan: 15.2 |
| **Category 2:**   - Adequate literacy in their own language* - Low-medium English proficiency (>10% low) - translated materials often available in NSW | Arabic | 161,688 | 15.9 | 43.9 | 19.4 | Common | 19.1 | 31.1 |
|  | Mandarin | 81,182 | 28.4 | 15.2 | 35.0 | Common | 46.4 | 28.7 |
| **Category 3:**   - Adequate literacy in their own language* - low-medium English proficiency (>10% low) - translated materials rare in NSW | Spanish | 28,243 | 11.8 | 25.4 | 18.8 | Uncommon | 30.8 | 20.7 |
|  | Croatian | 11,713 | 13.1 | 38.5 | 1.9 | Uncommon | 14.0 | 26.2 |
| **Category 4:**   - Low literacy in their own language* - low English proficiency - translated materials rare in NSW | Assyrian | 28,104 | 22.2 | 26.2 | 22.5 | Uncommon | 11.0 | 29.1 |
|  | Dinka | 1,622 | 16.7 | 25.9 | 10.7 | Uncommon | 8.9 | 38.3 |
|  | Dari | 24,553 | 24.9 | 23.1 | 26.1 | Uncommon | 16.6 | 31.2 |
|  | Khmer | 10,632 | 33.7 | 24.2 | 13.8 | Uncommon | 12.3 | 20.1 |

* Based on advice from New South Wales Local Health District staff (authors DZ, UT, YS, GV, TC)
