## Appendix 3 for "Main COVID-19 information sources in a culturally and linguistically diverse community in Sydney, Australia: A cross-sectional survey"

### COVID-19 Survey

#### Community Summary

### Arabic

A summary of our research into the views of the Arabic speaking community in Greater Western Sydney about COVID-19, conducted between 21/03/21 and 09/07/21

Health Literacy Hub

Sydney Health Literacy Lab

- Average 2.3/10 score for risk perception
- 65% would say Yes to a vaccine
- 83% would get tested 'No matter what'
- Top source for COVID-19 information: social media (68%)

- 80 people who speak Arabic as their main language at home took part
- 84% speak English very well/well (67 out of 80)
- 81% read Arabic very well/well (65 out of 80)
- 69% adequate health literacy (55 out of 80)

Country of birth

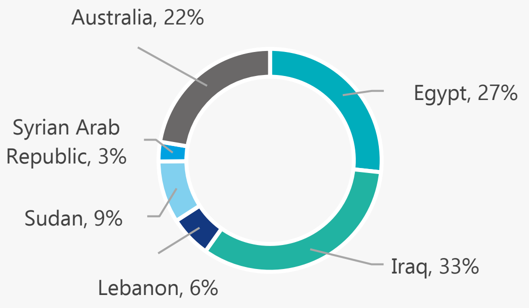


Age and gender

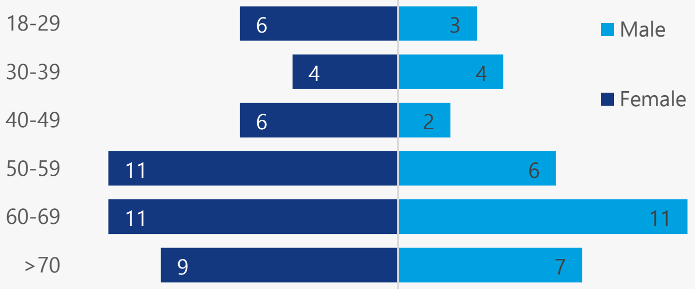


#### Top Sources for COVID-19 Information

1. **Social media (68%):** Facebook (82%); YouTube (44%); WhatsApp (40%)
2. **Official Australian source (49%):** Health professional (64%); Australian government websites (61%); Australian public TV (36%)
3. **Australian commercial source (49%):** Australian commercial TV (74%); Australian news/magazine website (29%)

**36%** get information about COVID-19 in a language other than English

Average of **2.8** out of 10 for difficulty finding COVID-19 information in Arabic that is easy to understand*

Average of **3.8** out of 10 for difficulty finding information in English that is easy to understand*

**where 10 = extremely difficult*

#### Top barriers to getting a COVID-19 vaccine

**65%**

If a COVID-19 vaccine is recommended to me, I will get it

**47%**

I am worried about side effects

**9%**

I need more information to make a decision

**10%**

I think the vaccine may not work well

**8%**

I do not think the vaccine will be safe

- **Risk perception very low (average of 2.3 out of 10)**
- **High intentions to perform COVID-19 prevention behaviours (average of 4.7 out of 5)**

On a scale of 0 to 10, how serious a public health problem do you think COVID-19 is currently, in Australia?

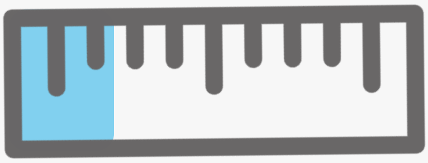

Where 1 is strongly disagree and 5 is strongly agree,
In the next 4 weeks, I will...

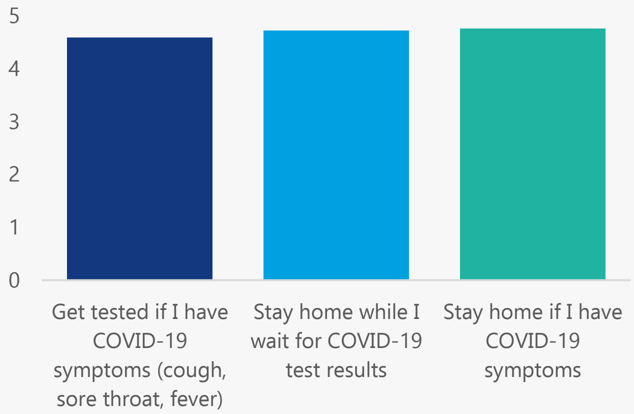


#### Top barriers to getting tested for COVID-19

**26%**

Testing is painful

**14%**

I already had a negative test so I don't need another one

**16%**

I don't know how, when and where to get tested

**9%**

I'm worried the results will be on my health record

**81%**

I will get tested no matter what

#### Impacts of COVID-19: **Employment**

- **36%** said their employment had changed as a result of COVID-19
- **16%** said they were ‘Not at all’ or ‘A little bit’ able to meet their **weekly expenses. 51%** were ‘Somewhat’ able to, and **35%** said ‘Quite a bit’ or ‘Very much’
- **43%** said they were ‘Not at all’ or ‘A little bit’ worried about the **financial problems** they will have in the future as a result of the pandemic. **34%** were ‘Somewhat’ worried, and **24%** said that they were ‘Quite a bit’ or ‘Very much’ worried

How has your employment changed?

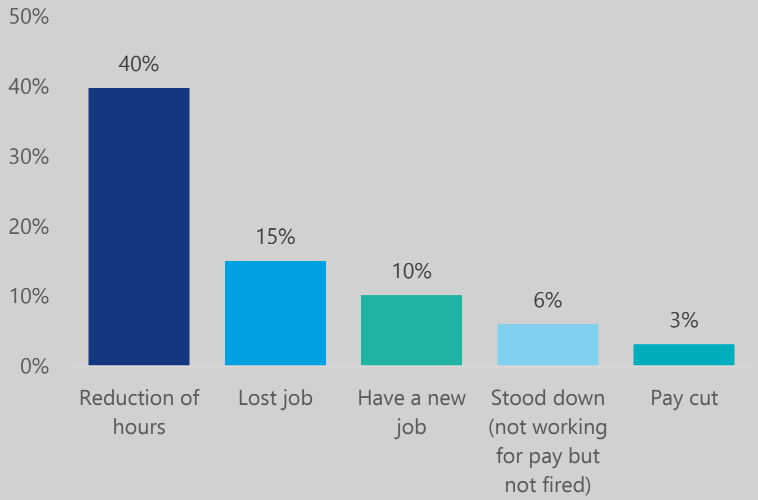


Where 1 is strongly disagree and 5 is strongly agree, Since the pandemic started...

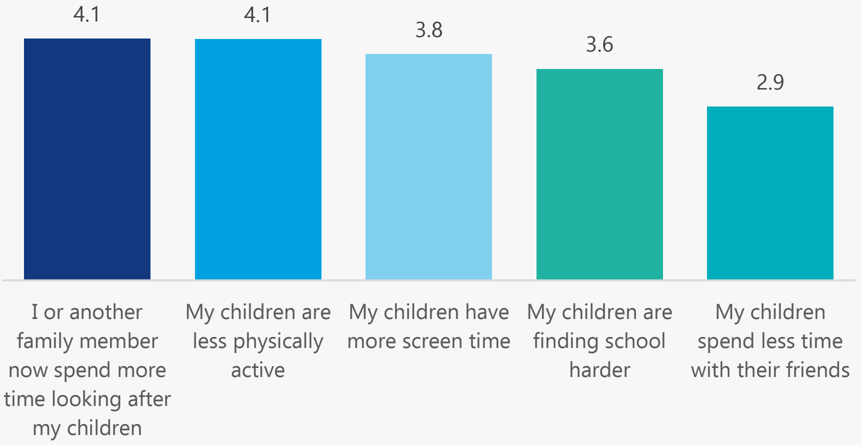


#### Impacts of COVID-19: **Relationships + Children**

- **80%** said COVID-19 has had no effect on their relationship with their partner
- **14%** said COVID-19 has had positive effects on their relationship with their partner
- **7%** said COVID-19 has had negative effects on their relationship with their partner

#### Impacts of COVID-19: **Mental Health**

Over the past week, how often have you felt nervous or "stressed" because of COVID-19?

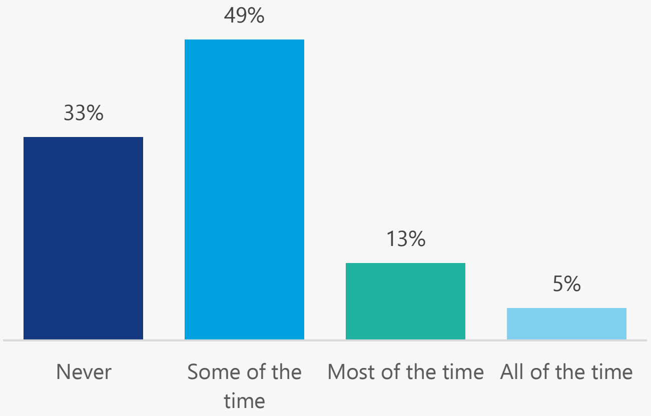


Over the past week, how often have you felt alone or lonely because of COVID-19?

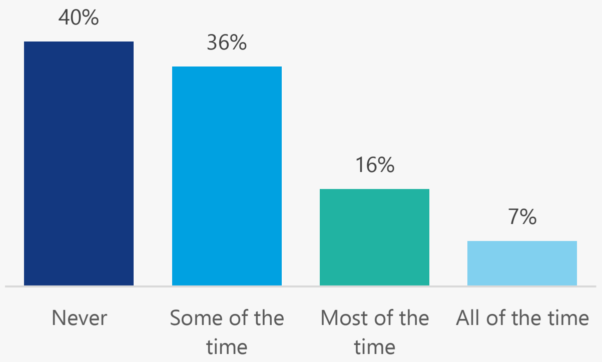


This research was a collaboration between the Sydney Health Literacy Lab (The University of Sydney), Health Literacy Hub (Western Sydney Local Health District), and Western Sydney, South Western Sydney, and Nepean Blue Mountains Local Health Districts. We would like to acknowledge and thank all staff, community members, and participants who made this project possible.

Health Literacy Hub

Sydney Health Literacy Lab

**Notes**

Raw counts are shown in the age and gender figure on page 1. Elsewhere in the report, counts were re-weighted to reflect the population profile in Greater Western Sydney.
This data was collected between 21st March and 9th July 2021. At the time of writing (2nd August 2021), the most recent outbreak commenced on 17th June 2020, and reached 448 cases by day 23 (9th July) when the survey closed.

You can find the University of Sydney Health Literacy Lab at https://sydneyhealthliteracylab.org.au/
The Western Sydney Health Literacy Hub can be found at https://www.healthliteracyhub.org.au/
You can find COVID-19 community resources for Western Sydney LHD at https://www.wslhd.health.nsw.gov.au/
Resources for South Western Sydney LHD at https://www.swslhd.health.nsw.gov.au/
And resources for Nepean Blue Mountains LHD at https://www.nbmlhd.health.nsw.gov.au/

This community summary was prepared by Carys Batcup. To reference the summary, please use this reference:
Ayre J, Muscat DMM, Mac O, Batcup C, Cvejic E, Pickles K, Dolan H, Bonner C, Mouwad D, Zachariah D, Turalic U, Santalucia Y, Chen T, Vasic G, McCaffery K. 2021. COVID-19 Survey Community Summary: Arabic. Available from http://healthliteracyhub.org.au

### COVID-19 Survey

#### Community Summary

### Assyrian

A summary of our research into the views of the Assyrian speaking community in Greater Western Sydney about COVID-19, conducted between 21/03/21 and 09/07/21

- Average 3.1/10 score for risk perception
- 41% would say Yes to a vaccine
- 80% would get tested 'No matter what'
- Top source for COVID-19 information was Australian commercial sources (63%)

- 133 people who speak Assyrian as their main language at home took part
- 68% speak English very well/well (90 out of 133)
- 61% read Assyrian very well/well (81 out of 133)
- 54% adequate health literacy (72 out of 133)

Country of birth

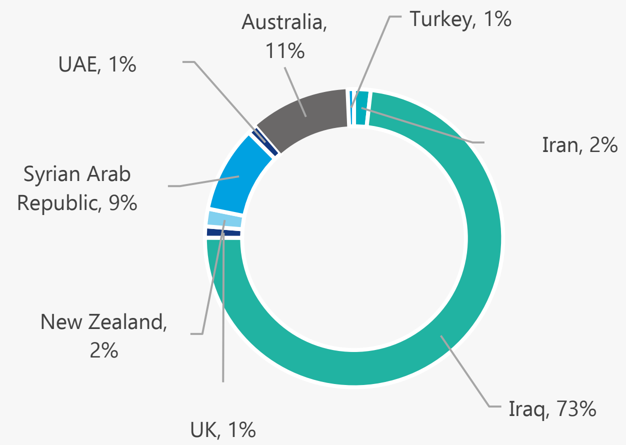


Age and gender

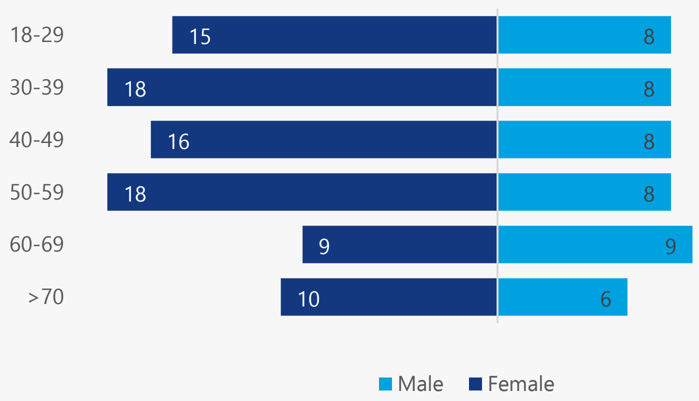


#### Top Sources for COVID-19 Information

1. **Australian commercial source (63%):** Australian commercial TV (84%); Australian news/magazine website (14%)
2. **Official Australian source (58%):** Australian public TV (73%); Health professional (52%); Australian government websites (28%)
3. **Social media (41%):** Facebook (94%); YouTube (39%); Instagram (30%)
4. **Friends or family living in Australia (32%):** living in Australia longer than the participant (88%)

**33%** get information about COVID-19 in a language other than English

Average of **5.6** out of 10 for difficulty finding COVID-19 information in Assyrian that is easy to understand*

Average of **4.2** out of 10 for difficulty finding information in English that is easy to understand*

**where 10 = extremely difficult*

#### Top barriers to getting a COVID-19 vaccine

**41%**

If a COVID-19 vaccine is recommended to me, I will get it

**29%**

I am worried about side effects

**14%**

I do not trust the drug companies

**20%**

I do not think the vaccine will be safe

**10%**

I am worried about how the vaccine may affect my other illness

- **Risk perception very low (average of 3.1 out of 10)**
- **High intentions to perform COVID-19 prevention behaviours (average of 4.5 out of 5)**

On a scale of 0 to 10, how serious a public health problem do you think COVID-19 is currently, in Australia?

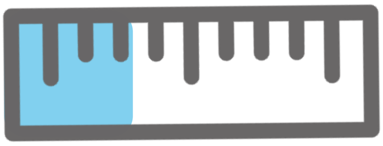

Where 1 is strongly disagree and 5 is strongly agree,
In the next 4 weeks, I will...

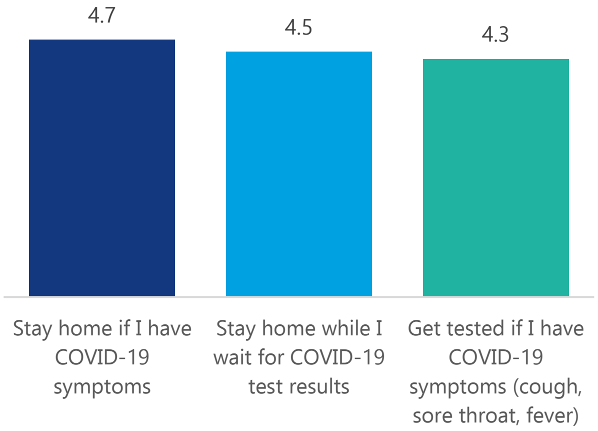


#### Top barriers to getting tested for COVID-19

**33%**

I'm worried I will get infected with COVID-19 at the testing clinic

**9%**

I'm worried about what others think of me

**25%**

Testing is painful

**9%**

I don't know how, when and where to get tested

**80%**

I will get tested no matter what

#### Impacts of COVID-19: **Employment**

- **18%** said their employment had changed as a result of COVID-19
- **29%** said they were ‘Not at all’ or ‘A little bit’ able to meet their **weekly expenses.** **29%** were ‘Somewhat’ able to, and **42%** said ‘Quite a bit’ or ‘Very much’
- **50%** said they were ‘Not at all’ or ‘A little bit’ worried about the **financial problems** they will have in the future as a result of the pandemic. **21%** were ‘Somewhat’ worried, and **29%** said that they were ‘Quite a bit’ or ‘Very much’ worried

How has your employment changed?

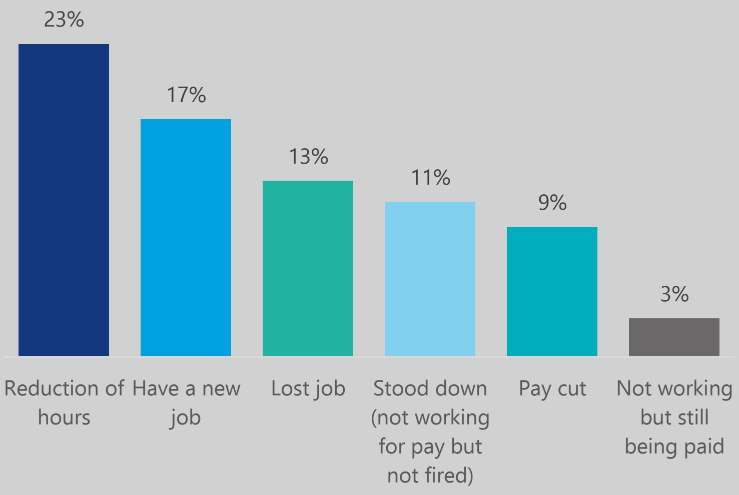


#### Impacts of COVID-19: **Relationships + Children**

Where 1 is strongly disagree and 5 is strongly agree, Since the pandemic started...

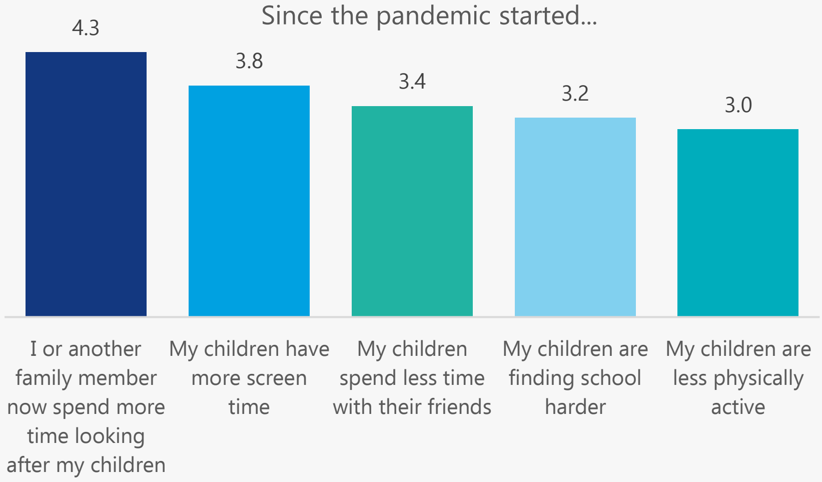


- **88%** said COVID-19 has had no effect on their relationship with their partner
- **10%** said COVID-19 has had positive effects on their relationship with their partner
- **3%** said COVID-19 has had negative effects on their relationship with their partner

#### Impacts of COVID-19: **Mental Health**

Over the past week, how often have you felt alone or lonely because of COVID-19?

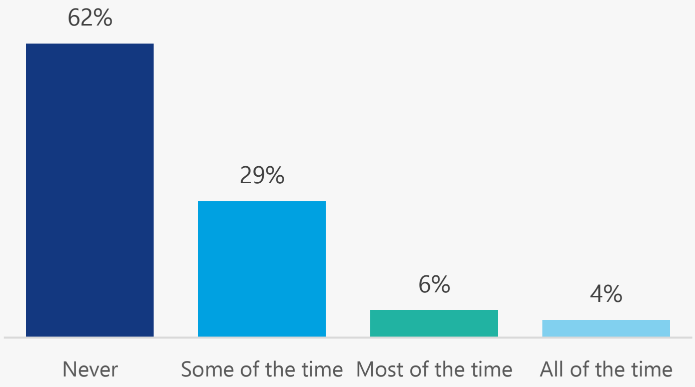


Over the past week, how often have you felt nervous or "stressed" because of COVID-19?

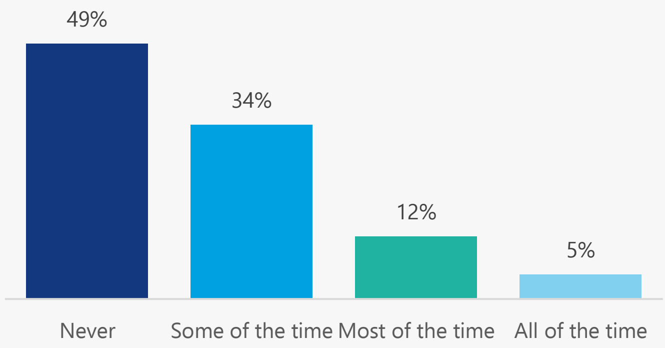


This research was a collaboration between the Sydney Health Literacy Lab (The University of Sydney), Health Literacy Hub (Western Sydney Local Health District), and Western Sydney, South Western Sydney, and Nepean Blue Mountains Local Health Districts. We would like to acknowledge and thank all staff, community members, and participants who made this project possible.

Health Literacy Hub

Sydney Health Literacy Lab

**Notes**

Raw counts are shown in the age and gender figure on page 1. Elsewhere in the report, counts were re-weighted to reflect the population profile in Greater Western Sydney.
This data was collected between 21st March and 9th July 2021. At the time of writing (2nd August 2021), the most recent outbreak commenced on 17th June 2020, and reached 448 cases by day 23 (9th July) when the survey closed.

You can find the University of Sydney Health Literacy Lab at https://sydneyhealthliteracylab.org.au/
The Western Sydney Health Literacy Hub can be found at https://www.healthliteracyhub.org.au/
You can find COVID-19 community resources for Western Sydney LHD at https://www.wslhd.health.nsw.gov.au/
Resources for South Western Sydney LHD at https://www.swslhd.health.nsw.gov.au/
And resources for Nepean Blue Mountains LHD at https://www.nbmlhd.health.nsw.gov.au/

This community summary was prepared by Carys Batcup. To reference the summary, please use this reference:
Ayre J, Muscat DMM, Mac O, Batcup C, Cvejic E, Pickles K, Dolan H, Bonner C, Mouwad D, Zachariah D, Turalic U, Santalucia Y, Chen T, Vasic G, McCaffery K. 2021. COVID-19 Survey Community Summary: Assyrian. Available from http://healthliteracyhub.org.au

### COVID-19 Survey

#### Community Summary

### Chinese

A summary of our research into the views of the Chinese speaking community in Greater Western Sydney about COVID-19, conducted between 21/03/21 and 09/07/21

Average 2.8/10 score for risk perception

46% would say Yes to a vaccine; main concerns were side effects + safety

74% would get tested 'No matter what'

Top source for COVID-19 information was social media (72%)

- 77 people who speak Chinese as their main language at home took part
- 58% speak English very well/well (45 out of 77)
- 96% read Chinese very well/well (74 out of 77)
- 52% adequate health literacy (40 out of 77)

Country of birth

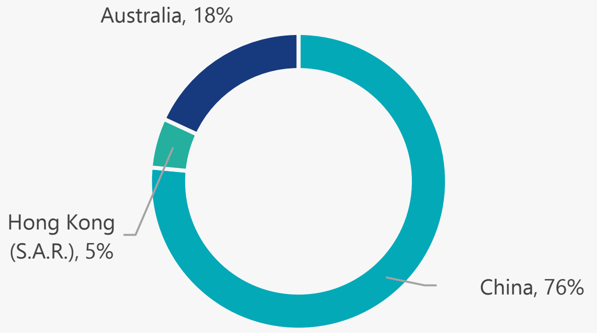


Age and gender

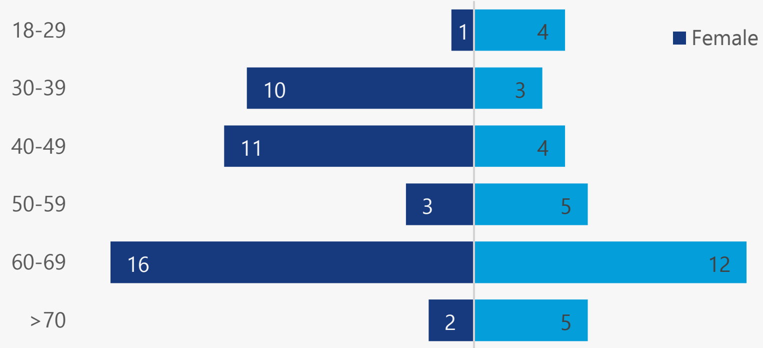


And one aged 30-39 who chose 'Prefer not to say'

##### Top Sources for COVID-19 Information

1. **Social media (72%):** WeChat (76%); YouTube (35%); Facebook (20%)
2. **Friends or family living in Australia (47%):** living in Australia longer than participant (78%)
3. **Australian commercial source (46%):** Australian news/magazine website (92%); Australian commercial TV (35%)
4. **Official Australian source (44%):** Australian government websites (63%); Australian public TV (50%)

**58%** get information about COVID-19 in a language other than English

Average of **3.4** out of 10 for difficulty finding COVID-19 information in Chinese that is easy to understand*

Average of **4.1** out of 10 for difficulty finding information in English that is easy to understand*

**where 10 = extremely difficult*

#### Top barriers to getting a COVID-19 vaccine

**46%**

If a COVID-19 vaccine is recommended to me, I will get it

**44%**

I am worried about side effects

**8%**

I need more information to make a decision

**29%**

I do not think the vaccine will be safe

**7%**

I think the vaccine may not work well

- **Risk perception very low (average of 2.8 out of 10)**
- **High intentions to perform COVID-19 prevention behaviours (average of 4.8 out of 5)**

On a scale of 0 to 10, how serious a public health problem do you think COVID-19 is currently, in Australia?

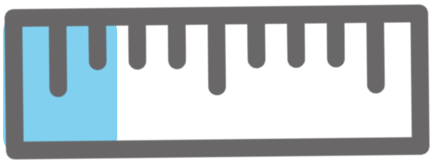

Where 1 is strongly disagree and 5 is strongly agree,
In the next 4 weeks, I will...

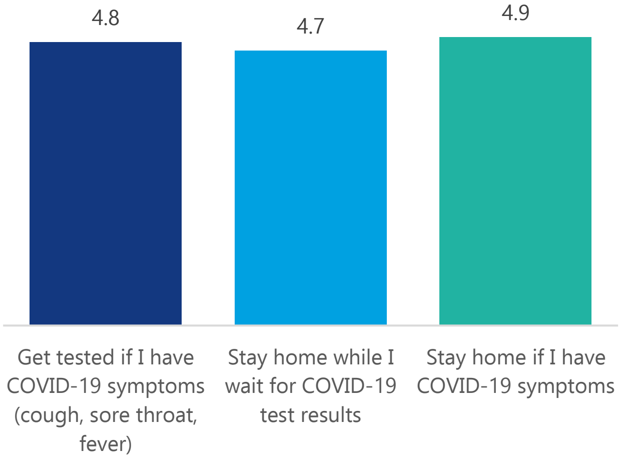


#### Top barriers to getting tested for COVID-19

**50%**

I'm worried I will get infected with COVID-19 at the testing clinic

**15%**

I already had a negative test so I don't need another one

**17%**

I don't know how, when and where to get tested

**6%**

Testing is painful

**74%**

I will get tested no matter what

#### Impacts of COVID-19: **Employment**

- **39%** said their employment had changed as a result of COVID-19
- **27%** said they were 'Not at all' or 'A little bit' able to meet their **weekly expenses. 23%** were 'Somewhat' able to, and **49%** said 'Quite a bit' or 'Very much'
- **56%** said they were 'Not at all' or 'A little bit' worried about the **financial problems** they will have in the future as a result of the pandemic. **24%** were 'Somewhat' worried, and **18%** said that they were 'Quite a bit' or 'Very much' worried

How has your employment changed?

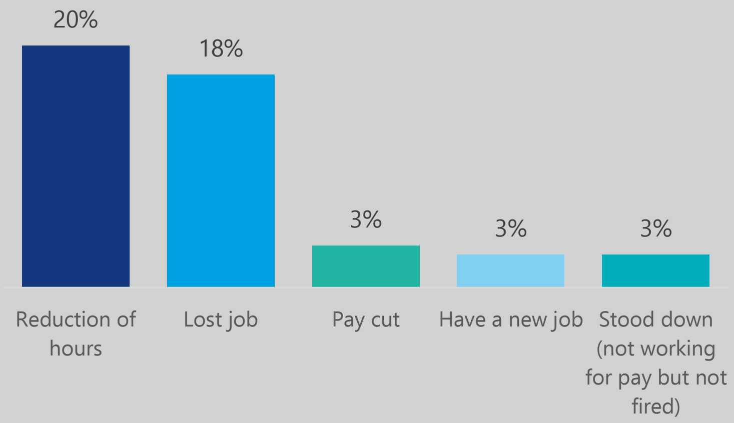


#### Impacts of COVID-19: **Relationships + Children**

Where 1 is strongly disagree and 5 is strongly agree, Since the pandemic started...

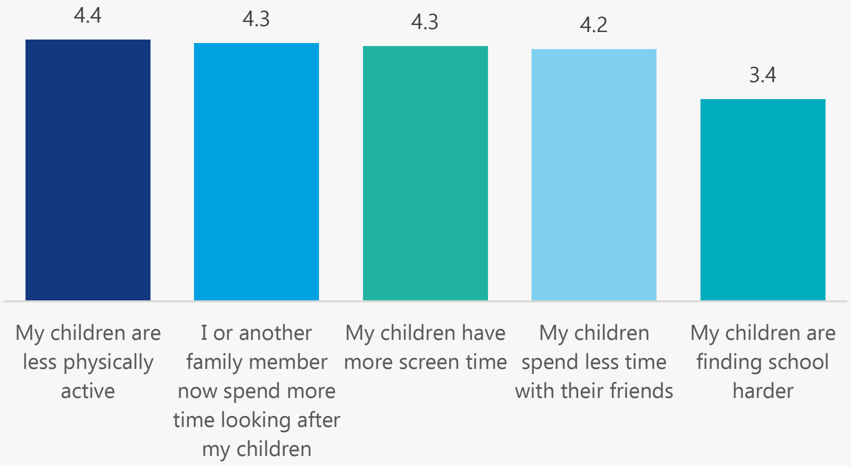


- **66%** said COVID-19 has had no effect on their relationship with their partner
- **20%** said COVID-19 has had some negative effects on their relationship with their partner
- **14%** said COVID-19 has had some positive effects on their relationship with their partner

#### Impacts of COVID-19: **Mental Health**

Over the past week, how often have you felt nervous or "stressed" because of COVID-19?

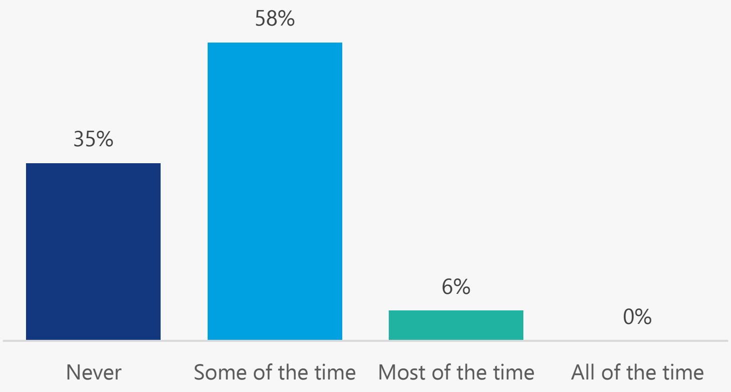


Over the past week, how often have you felt alone or lonely because of COVID-19?

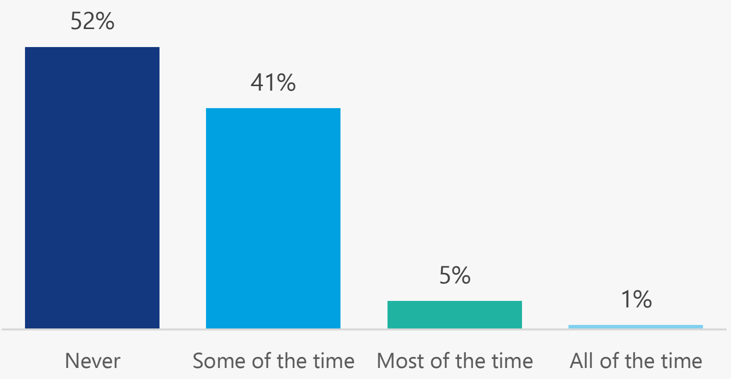


This research was a collaboration between the Sydney Health Literacy Lab (The University of Sydney), Health Literacy Hub (Western Sydney Local Health District), and Western Sydney, South Western Sydney, and Nepean Blue Mountains Local Health Districts. We would like to acknowledge and thank all staff, community members, and participants who made this project possible.

Health Literacy Hub

Sydney Health Literacy Lab

**Notes**

Raw counts are shown in the age and gender figure on page 1. Elsewhere in the report, counts were re-weighted to reflect the population profile in Greater Western Sydney.
This data was collected between 21st March and 9th July 2021. At the time of writing (2nd August 2021), the most recent outbreak commenced on 17th June 2020, and reached 448 cases by day 23 (9th July) when the survey closed.

You can find the University of Sydney Health Literacy Lab at https://sydneyhealthliteracylab.org.au/
The Western Sydney Health Literacy Hub can be found at https://www.healthliteracyhub.org.au/
You can find COVID-19 community resources for Western Sydney LHD at https://www.wslhd.health.nsw.gov.au/
Resources for South Western Sydney LHD at https://www.swslhd.health.nsw.gov.au/
And resources for Nepean Blue Mountains LHD at https://www.nbmlhd.health.nsw.gov.au/

This community summary was prepared by Carys Batcup. To reference the summary, please use this reference:
Ayre J, Muscat DMM, Mac O, Batcup C, Cvejic E, Pickles K, Dolan H, Bonner C, Mouwad D, Zachariah D, Turalic U, Santalucia Y, Chen T, Vasic G, McCaffery K. 2021. COVID-19 Survey Community Summary: Chinese. Available from http://healthliteracyhub.org.au

### COVID-19 Survey

#### Community Summary

### Croatian

A summary of our research into the views of the Croatian speaking community in Greater Western Sydney about COVID-19, conducted between 21/03/21 and 09/07/21

- Average 4.0/10 score for risk perception
- 43% would say Yes to a vaccine
- 59% would get tested 'No matter what'
- Top source for COVID-19 information was overseas sources (98%): mainly websites (82%)

- 121 people who speak Croatian as their main language at home took part
- 38% speak English very well/well (46 out of 121)
- 100% read Croatian very well/well (121 out of 121)
- 32% adequate health literacy (39 out of 121)

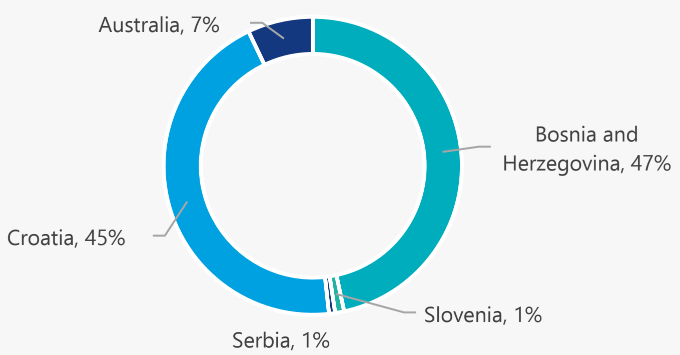


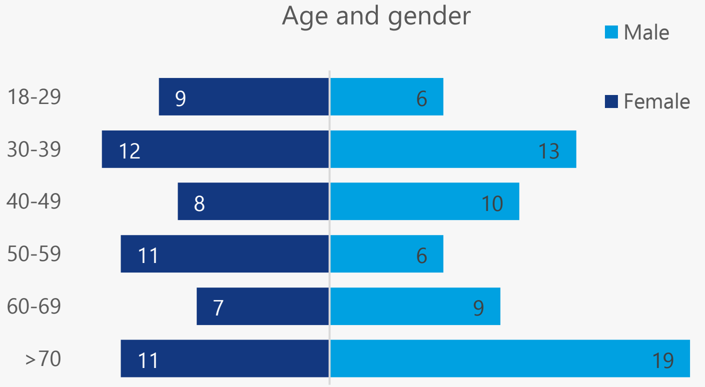


##### Top Sources for COVID-19 Information

1. **Overseas information sources (98%):** Overseas websites (82%); Overseas TV (74%); Family or friends living overseas (63%)
2. **Community (65%):** Community leader (69%); Community TV (34%); Religious leader (33%)
3. **Friends or family living in Australia (63%):** living in Australia same amount of time or less than the participant (95%)
4. **Official Australian source (60%):** Health professional (99%); Australian government websites (18%)

**98%** get information about COVID-19 in a language other than English

Average of **3.4** out of 10 for difficulty finding COVID-19 information in Croatian that is easy to understand*

Average of **6.6** out of 10 for difficulty finding information in English that is easy to understand*

**where 10 = extremely difficult*

##### Top barriers to getting a COVID-19 vaccine

**43%**

If a COVID-19 vaccine is recommended to me, I will get it

**60%**

I do not think the vaccine will be safe

**27%**

I am worried about the side effects

- **Risk perception low (average of 4.0 out of 10)**
- **High intentions to perform COVID-19 prevention behaviours (average of 4.3 out of 5)**

On a scale of 0 to 10, how serious a public health problem do you think COVID-19 is currently, in Australia?

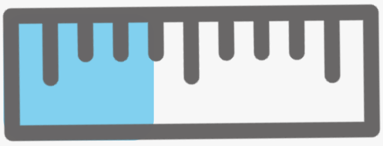

Where 1 is strongly disagree and 5 is strongly agree,
In the next 4 weeks, I will...

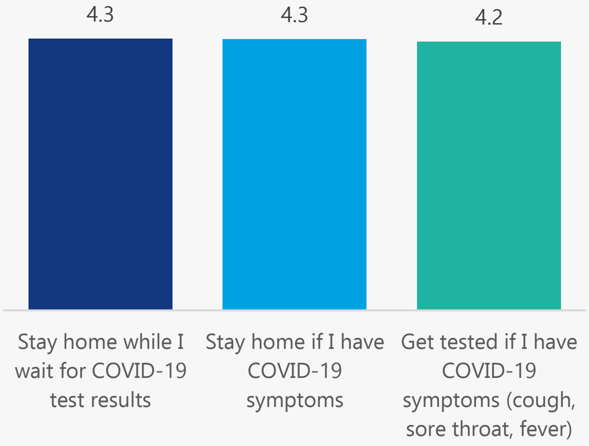


##### Top barriers to getting tested for COVID-19

**29%**

I'll forget to get tested

**13%**

I already had a negative test so I don't need another one

**22%**

I'm worried about what will happen to my visa if I test positive

**13%**

I'm worried the results will be on my health record

**59%**

I will get tested no matter what

#### Impacts of COVID-19: **Employment**

- **42%** said their employment had changed as a result of COVID-19
- **4%** said they were ‘Not at all’ or ‘A little bit’ able to meet their **weekly expenses. 34%** were ‘Somewhat’ able to, and **62%** said ‘Quite a bit’ or ‘Very much’
- **32%** said they were ‘Not at all’ or ‘A little bit’ worried about the **financial problems** they will have in the future as a result of the pandemic. **22%** were ‘Somewhat’ worried, and **47%** said that they were ‘Quite a bit’ or ‘Very much’ worried

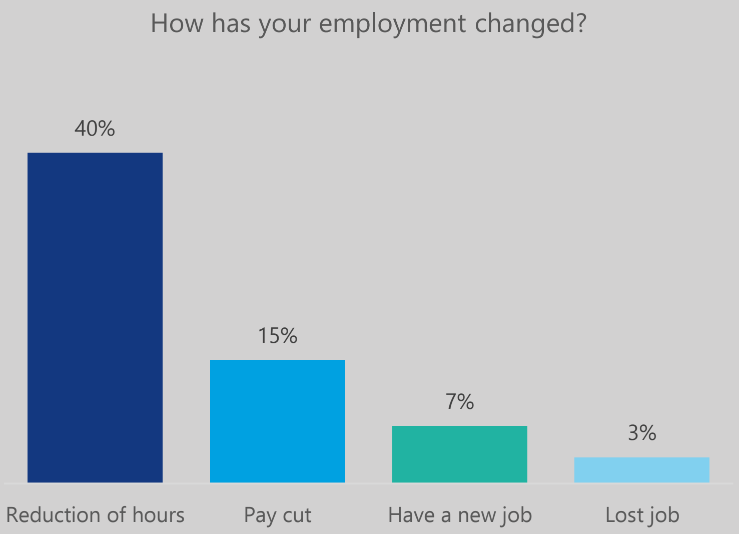


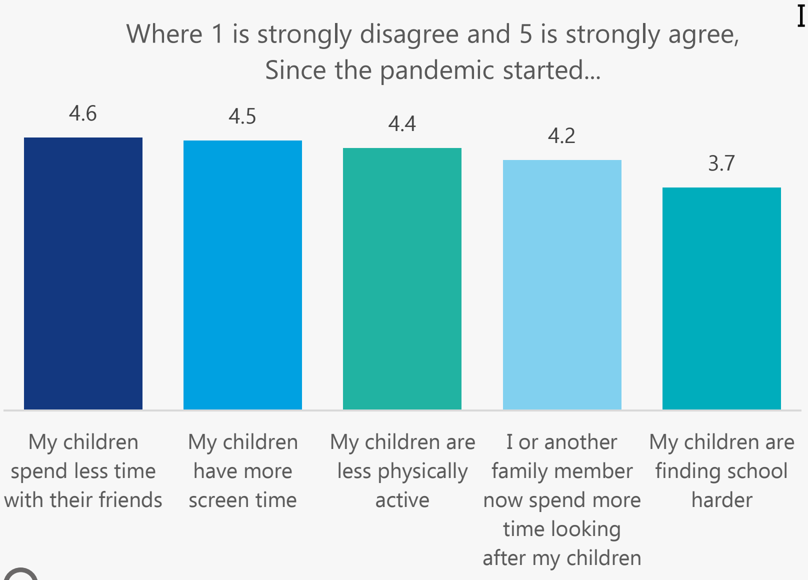


#### Impacts of COVID-19: **Relationships + Children**

- **46%** said COVID-19 has had no effect on their relationship with their partner
- **38%** said COVID-19 has had negative effects on their relationship with their partner
- **16%** said COVID-19 has had positive effects on their relationship with their partner

#### Impacts of COVID-19: **Mental Health**

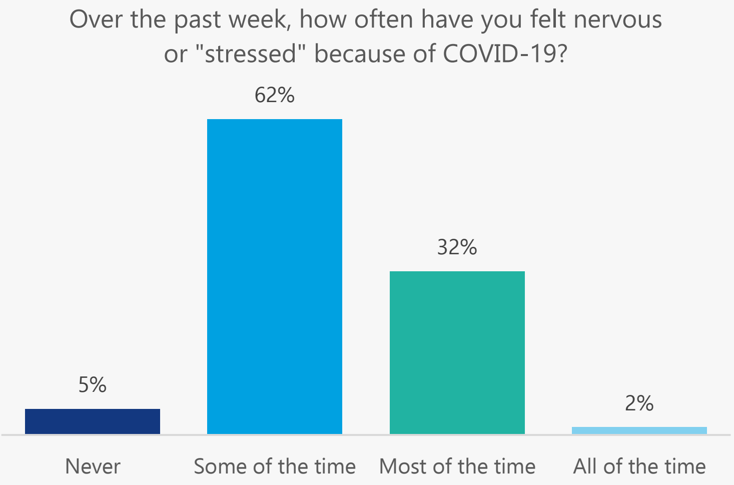


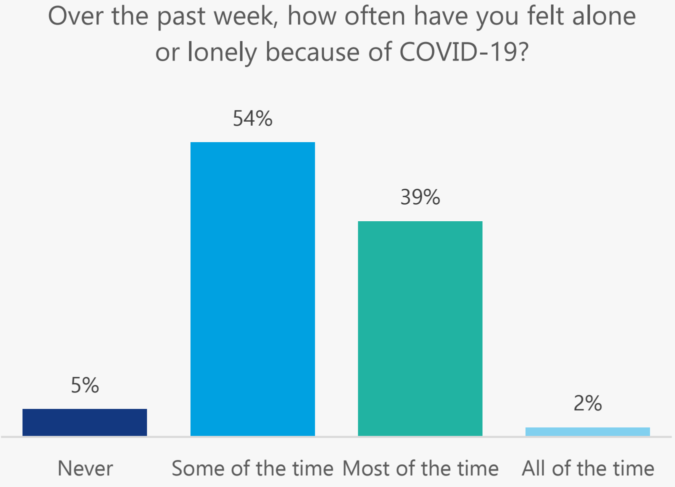


This research was a collaboration between the Sydney Health Literacy Lab (The University of Sydney), Health Literacy Hub (Western Sydney Local Health District), and Western Sydney, South Western Sydney, and Nepean Blue Mountains Local Health Districts. We would like to acknowledge and thank all staff, community members, and participants who made this project possible.

Health Literacy Hub

Sydney Health Literacy Lab

**Notes**

Raw counts are shown in the age and gender figure on page 1. Elsewhere in the report, counts were re-weighted to reflect the population profile in Greater Western Sydney.

This data was collected between 21st March and 9th July 2021. At the time of writing (2nd August 2021), the most recent outbreak commenced on 17th June 2020, and reached 448 cases by day 23 (9th July) when the survey closed.

You can find the University of Sydney Health Literacy Lab at https://sydneyhealthliteracylab.org.au/
The Western Sydney Health Literacy Hub can be found at https://www.healthliteracyhub.org.au/
You can find COVID-19 community resources for Western Sydney LHD at https://www.wslhd.health.nsw.gov.au/
Resources for South Western Sydney LHD at https://www.swslhd.health.nsw.gov.au/
And resources for Nepean Blue Mountains LHD at https://www.nbmlhd.health.nsw.gov.au/

This community summary was prepared by Carys Batcup. To reference the summary, please use this reference:
Ayre J, Muscat DMM, Mac O, Batcup C, Cvejic E, Pickles K, Dolan H, Bonner C, Mouwad D, Zachariah D, Turalic U, Santalucia Y, Chen T, Vasic G, McCaffery K. 2021. COVID-19 Survey Community Summary: Croatian. Available from http://healthliteracyhub.org.au

### COVID-19 Survey

#### Community Summary

### Dari

A summary of our research into the views of the Dari speaking community in Greater Western Sydney about COVID-19, conducted between 21/03/21 and 09/07/21

- Average 7.9/10 score for risk perception
- 34% would say Yes to a vaccine
- 74% would get tested 'No matter what'
- Top source for COVID-19 information was social media (73%)

- 44 people who speak Dari as their main language at home took part
- 59% speak English very well/well (26 out of 44)
- 92% read Dari very well/well (40 out of 44)
- 35% adequate health literacy (15 out of 44)

Country of birth

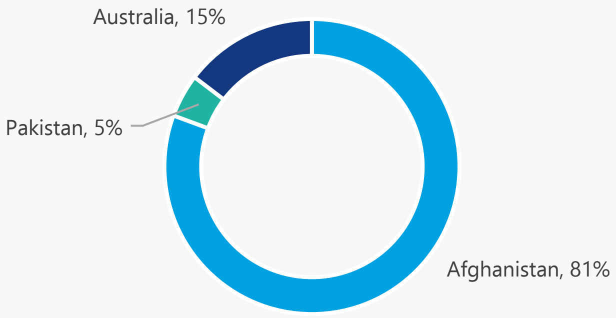


Age and gender

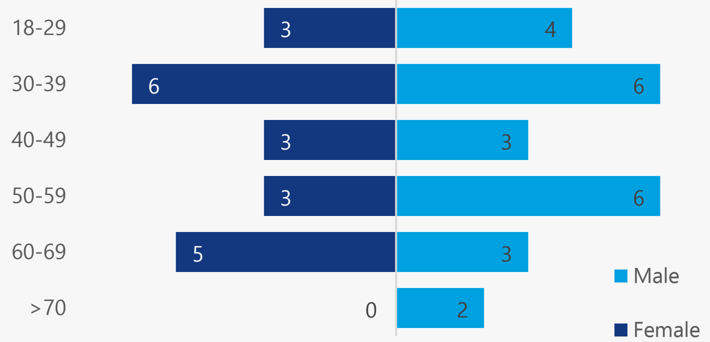


##### Top Sources for COVID-19 Information

1. **Social media (73%):** Facebook (97%); YouTube (67%); Instagram (36%)
2. **Official Australian source (67%):** Australian public TV (47%); Health professional (39%)
3. **Friends or family living in Australia (54%):** living in Australia more years than the participant (93%); living in Australia the same or fewer years than the participant (81%)
4. **Australian commercial source (39%):** Australian commercial TV (84%); Australian news/magazine website (37%)

**58%** get information about COVID-19 in a language other than English

Average of **7.0** out of 10 for difficulty finding COVID-19 information in Dari that is easy to understand*

Average of **7.0** out of 10 for difficulty finding information in English that is easy to understand*

**where 10 = extremely difficult*

#### Top barriers to getting a COVID-19 vaccine

**34%**

If a COVID-19 vaccine is recommended to me, I will get it

I want to wait so that I can see how other countries go first

**38%**

**26%**

I am worried about side effects

I am worried about how the vaccine may affect my other illness

**12%**

**11%**

I need more information to make a decision

- **Risk perception high (average of 7.9 out of 10)**
- **High intentions to perform COVID-19 prevention behaviours (average of 4.8 out of 5)**

On a scale of 0 to 10, how serious a public health problem do you think COVID-19 is currently, in Australia?

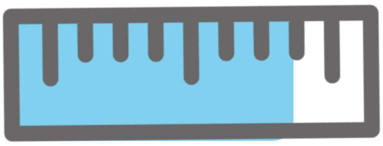

Where 1 is strongly disagree and 5 is strongly agree,
In the next 4 weeks, I will...

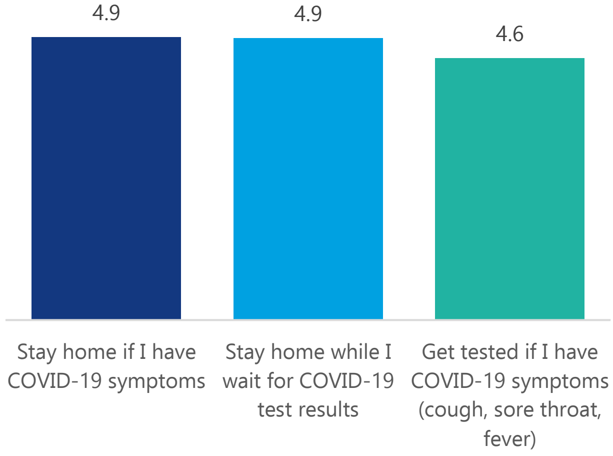


#### Top barriers to getting tested for COVID-19

**35%**

I don't know how, when and where to get tested

**13%**

It's too difficult or expensive to get tested

**21%**

I'm worried about what will happen to my visa if I test positive

**12%**

I'm worried I will get infected with COVID-19 at the testing clinic

**74%**

I will get tested no matter what

#### Impacts of COVID-19: **Employment**

- **57%** said their employment had changed as a result of COVID-19
- **19%** said they were ‘Not at all’ or ‘A little bit’ able to meet their **weekly expenses. 55%** were ‘Somewhat’ able to, and **26%** said ‘Quite a bit’ or ‘Very much’
- **7%** said they were ‘Not at all’ or ‘A little bit’ worried about the **financial problems** they will have in the future as a result of the pandemic. **33%** were ‘Somewhat’ worried, and **60%** said that they were ‘Quite a bit’ or ‘Very much’ worried

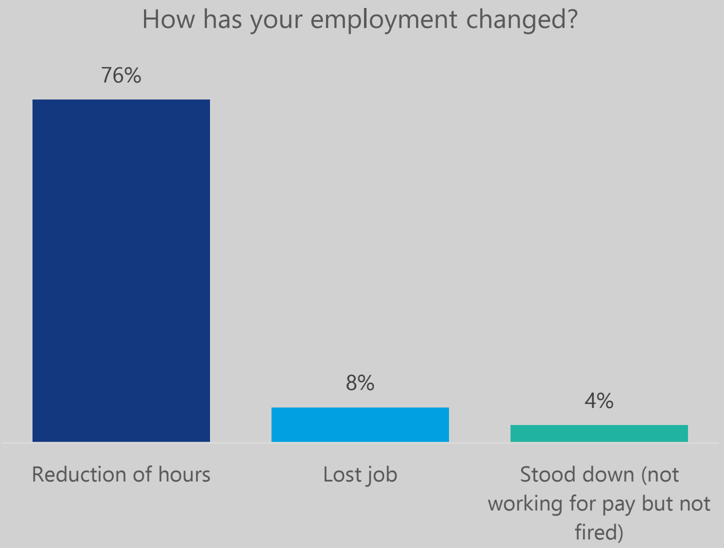


#### Impacts of COVID-19: **Relationships + Children**

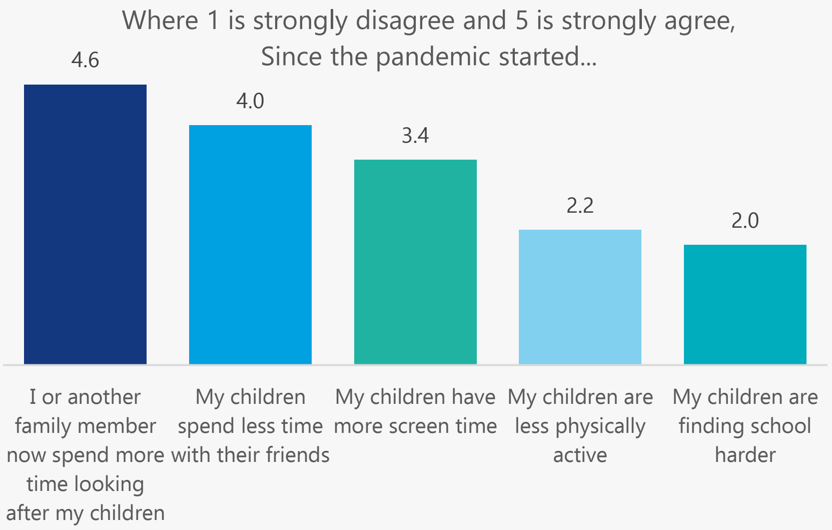


- **60%** said COVID-19 has had no effect on their relationship with their partner
- **33%** said COVID-19 has had negative effects on their relationship with their partner
- **7%** said COVID-19 has had positive effects on their relationship with their partner

#### Impacts of COVID-19: **Mental Health**

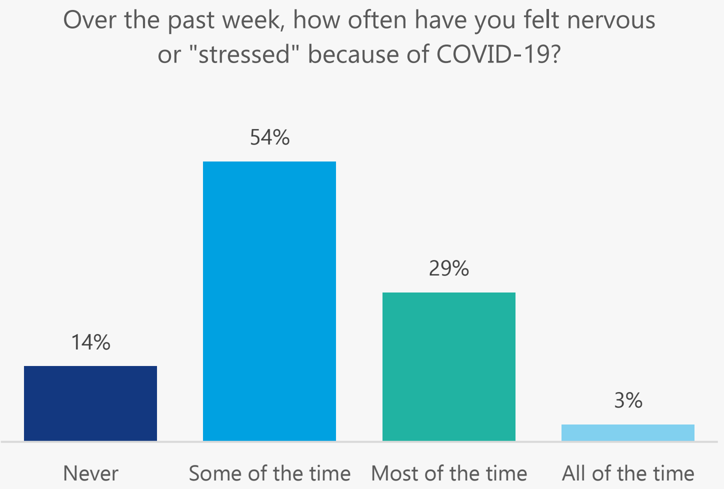


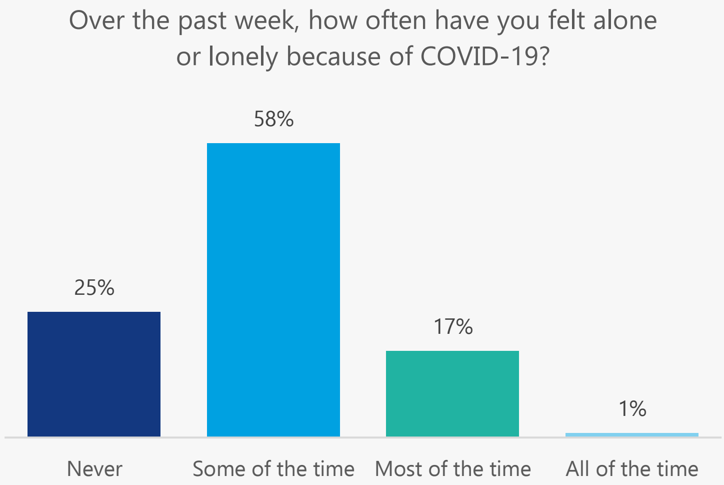


This research was a collaboration between the Sydney Health Literacy Lab (The University of Sydney), Health Literacy Hub (Western Sydney Local Health District), and Western Sydney, South Western Sydney, and Nepean Blue Mountains Local Health Districts. We would like to acknowledge and thank all staff, community members, and participants who made this project possible.

Health Literacy Hub

Sydney Health Literacy Lab

**Notes**

Raw counts are shown in the age and gender figure on page 1. Elsewhere in the report, counts were re-weighted to reflect the population profile in Greater Western Sydney.
This data was collected between 21st March and 9th July 2021. At the time of writing (2nd August 2021), the most recent outbreak commenced on 17th June 2020, and reached 448 cases by day 23 (9th July) when the survey closed.

You can find the University of Sydney Health Literacy Lab at https://sydneyhealthliteracylab.org.au/
The Western Sydney Health Literacy Hub can be found at https://www.healthliteracyhub.org.au/
You can find COVID-19 community resources for Western Sydney LHD at https://www.wslhd.health.nsw.gov.au/
Resources for South Western Sydney LHD at https://www.swslhd.health.nsw.gov.au/
And resources for Nepean Blue Mountains LHD at https://www.nbmlhd.health.nsw.gov.au/

This community summary was prepared by Carys Batcup. To reference the summary, please use this reference:
Ayre J, Muscat DMM, Mac O, Batcup C, Cvejic E, Pickles K, Dolan H, Bonner C, Mouwad D, Zachariah D, Turalic U, Santalucia Y, Chen T, Vasic G, McCaffery K. 2021. COVID-19 Survey Community Summary: Dari. Available from http://healthliteracyhub.org.au

### COVID-19 Survey

#### Community Summary

### Dinka

A summary of our research into the views of the Dinka speaking community in Greater Western Sydney about COVID-19, conducted between 21/03/21 and 09/07/21

- Average 5.5/10 score for risk perception
- 49% would say Yes to a vaccine: main barrier is worry about side effects
- 74% would get tested 'No matter what'
- Top source for COVID-19 information was official Australian source (67%)

- 63 people who speak Dinka as their main language at home took part
- 98% speak English very well/well (62 out of 63)
- 66% read Dinka very well/well (41 out of 63)
- 74% adequate health literacy (47 out of 63)

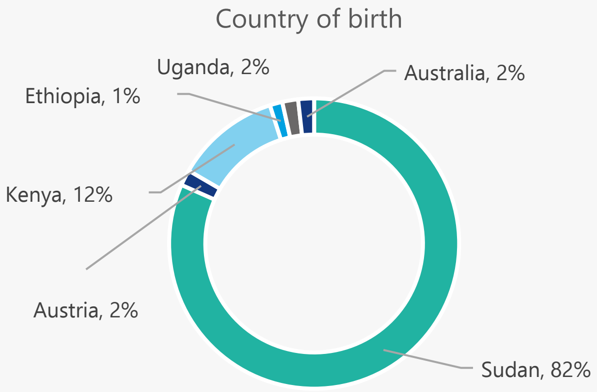


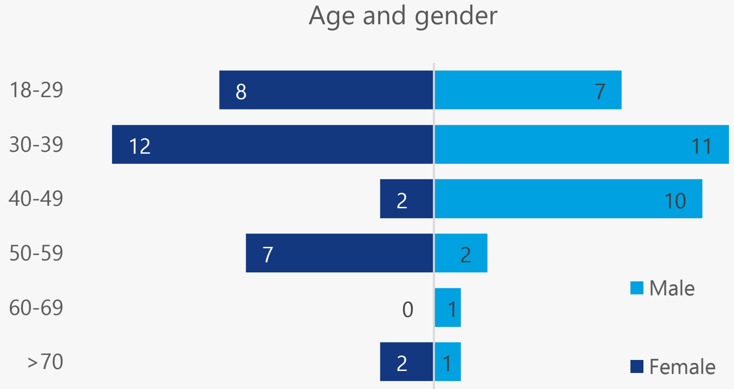


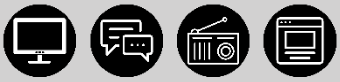

##### Top Sources for COVID-19 Information

1. **Official Australian source (67%):** Australian public TV (91%); Health professional (42%); Australian government websites (27%); Australian public radio or podcasts (21%)
2. **Social media (65%):** Facebook (85%); YouTube (39%); Instagram (36%)
3. **Australian commercial source (55%):** Australian commercial TV (87%); Australian news/magazine website (24%)

**5%** get information about COVID-19 in a language other than English

Average of **5.6** out of 10 for difficulty finding COVID-19 information in Dinka that is easy to understand*

Average of **2.6** out of 10 for difficulty finding information in English that is easy to understand*

**where 10 = extremely difficult*

**Top barriers to getting a COVID-19 vaccine**

**49%**

If a COVID-19 vaccine is recommended to me, I will get it

**32%**

I am worried about side effects

**11%**

I do not think the vaccine will be safe

**17%**

I need more information to make a decision

**9%**

I do not trust the drug companies

- **Risk perception medium (average of 5.5 out of 10)**
- **High intentions to perform COVID-19 prevention behaviours (average of 4.7 out of 5)**

On a scale of 0 to 10, how serious a public health problem do you think COVID-19 is currently, in Australia?

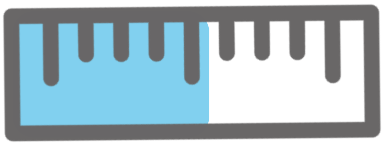

Where 1 is strongly disagree and 5 is strongly agree,
In the next 4 weeks, I will...

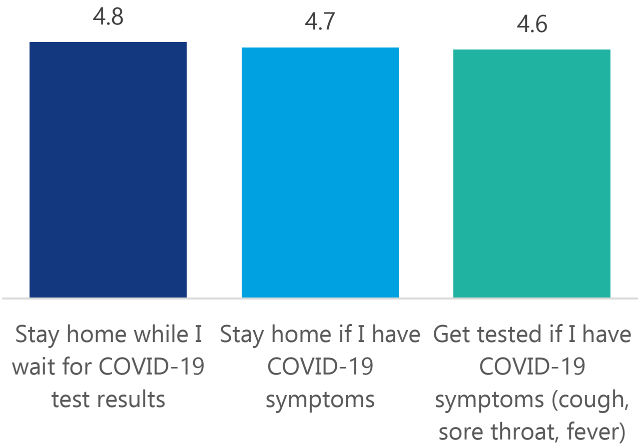


**Top barriers to getting tested for COVID-19**

**32%**

Testing is painful

**15%**

I'll forget to get tested

**19%**

I'm worried I will get infected with COVID-19 at the testing clinic

**9%**

I already had a negative test so I don't need another one

**74%**

I will get tested no matter what

#### Impacts of COVID-19: **Employment**

- **40%** said their employment had changed as a result of COVID-19
- **24%** said they were 'Not at all' or 'A little bit' able to meet their **weekly expenses.** **16%** were 'Somewhat' able to, and **59%** said 'Quite a bit' or 'Very much'
- **23%** said they were 'Not at all' or 'A little bit' worried about the **financial problems** they will have in the future as a result of the pandemic. **15%** were 'Somewhat' worried, and **62%** said that they were 'Quite a bit' or 'Very much' worried

How has your employment changed?

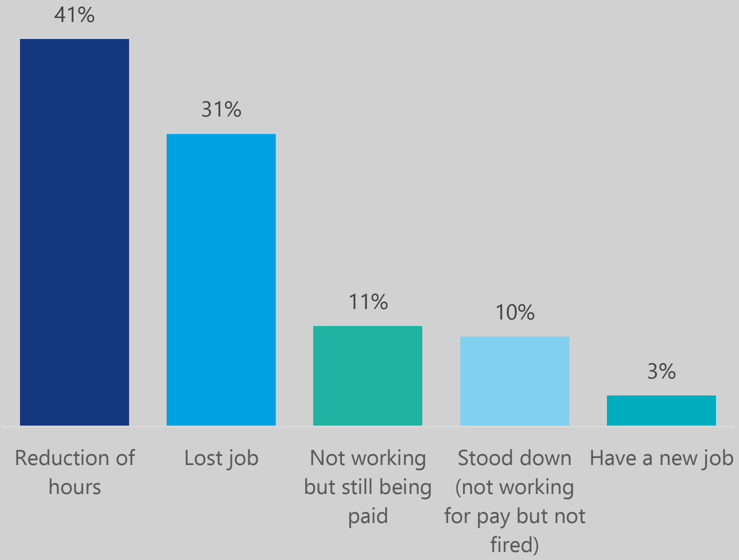


#### Impacts of COVID-19: **Relationships + Children**

Where 1 is strongly disagree and 5 is strongly agree, Since the pandemic started...

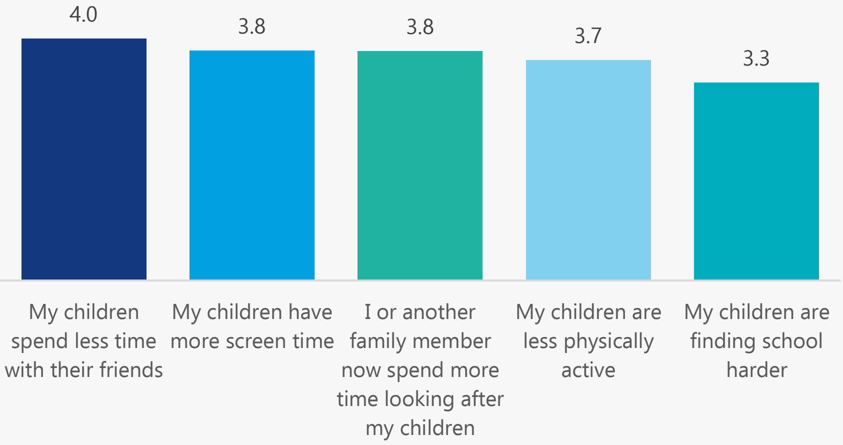


- **70%** said COVID-19 has had no effect on their relationship with their partner
- **19%** said COVID-19 has had negative effects on their relationship with their partner
- **11%** said COVID-19 has had positive effects on their relationship with their partner

#### Impacts of COVID-19: **Mental Health**

Over the past week, how often have you felt nervous or "stressed" because of COVID-19?

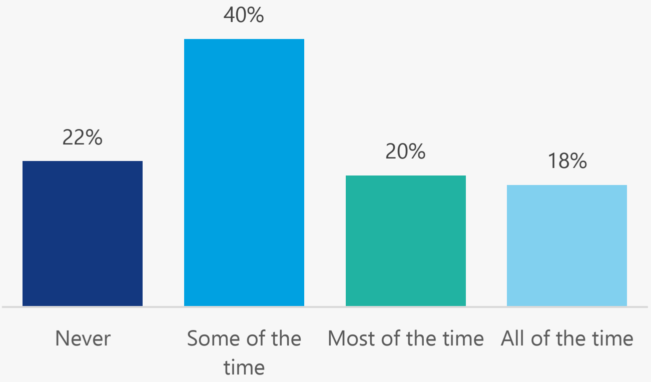


Over the past week, how often have you felt alone or lonely because of COVID-19?

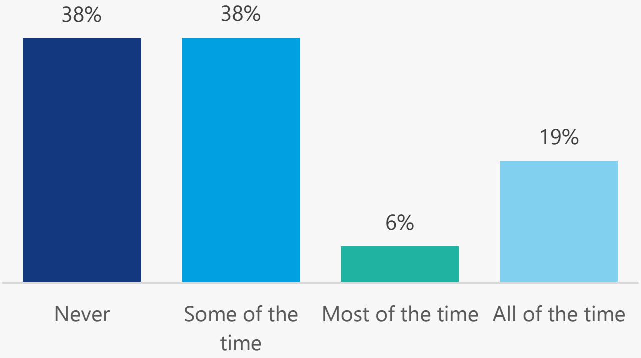


This research was a collaboration between the Sydney Health Literacy Lab (The University of Sydney), Health Literacy Hub (Western Sydney Local Health District), and Western Sydney, South Western Sydney, and Nepean Blue Mountains Local Health Districts. We would like to acknowledge and thank all staff, community members, and participants who made this project possible.

Health Literacy Hub

Sydney Health Literacy Lab

**Notes**

Raw counts are shown in the age and gender figure on page 1. Elsewhere in the report, counts were re-weighted to reflect the population profile in Greater Western Sydney.
This data was collected between 21st March and 9th July 2021. At the time of writing (2nd August 2021), the most recent outbreak commenced on 17th June 2020, and reached 448 cases by day 23 (9th July) when the survey closed.

You can find the University of Sydney Health Literacy Lab at https://sydneyhealthliteracylab.org.au/
The Western Sydney Health Literacy Hub can be found at https://www.healthliteracyhub.org.au/
You can find COVID-19 community resources for Western Sydney LHD at https://www.wslhd.health.nsw.gov.au/
Resources for South Western Sydney LHD at https://www.swslhd.health.nsw.gov.au/
And resources for Nepean Blue Mountains LHD at https://www.nbmlhd.health.nsw.gov.au/

This community summary was prepared by Carys Batcup. To reference the summary, please use this reference:
Ayre J, Muscat DMM, Mac O, Batcup C, Cvejic E, Pickles K, Dolan H, Bonner C, Mouwad D, Zachariah D, Turalic U, Santalucia Y, Chen T, Vasic G, McCaffery K. 2021. COVID-19 Survey Community Summary: Dinka. Available from http://healthliteracyhub.org.au

### COVID-19 Survey

#### Community Summary

### Hindi

A summary of our research into the views of the Hindi speaking community in Greater Western Sydney about COVID-19, conducted between 21/03/21 and 09/07/21

- Average 6.3/10 score for risk perception
- 82% would say Yes to a vaccine
- 99% would get tested 'No matter what'
- Top source for COVID-19 information was Australian commercial TV (68%)

- 42 people who speak Hindi as their main language at home took part
- 96% speak English very well/well (40 out of 42)
- 97% read Hindi very well/well (41 out of 42)
- 88% adequate health literacy (37 out of 42)

Country of birth

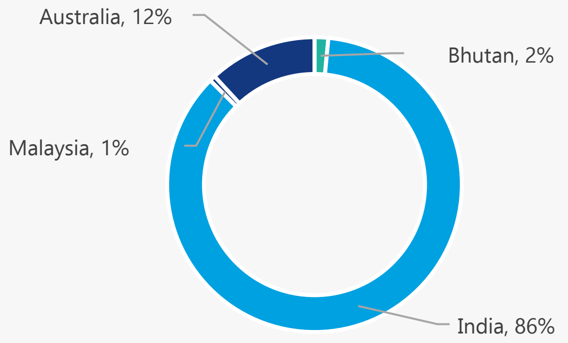


Age and gender

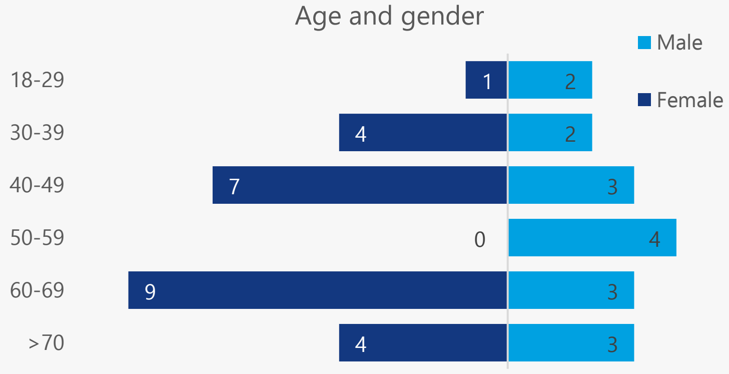


##### Top Sources for COVID-19 Information

1. **Australian commercial source (74%):** Australian commercial TV (92%)
2. **Social media (71%):** WhatsApp (65%); Facebook (63%); YouTube (50%); Instagram (42%)
3. **Official Australian source (69%):** Australian public TV (80%); Health professional (47%)
4. **Overseas information sources (33%):** Overseas TV (88%)

**15%** get information about COVID-19 in a language other than English

Average of **3.3** out of 10 for difficulty finding COVID-19 information in Hindi that is easy to understand*

Average of **1.7** out of 10 for difficulty finding information in English that is easy to understand*

**where 10 = extremely difficult*

#### Top barriers to getting a COVID-19 vaccine

**82%**

If a COVID-19 vaccine is recommended to me, I will get it

**62%**

I need more information to make a decision

**38%**

I am worried about the side effects

- **Risk perception medium (average of 6.3 out of 10)**
- **High intentions to perform COVID-19 prevention behaviours (average of 4.2 out of 5)**

On a scale of 0 to 10, how serious a public health problem do you think COVID-19 is currently, in Australia?

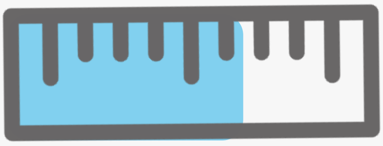

Where 1 is strongly disagree and 5 is strongly agree,
In the next 4 weeks, I will...

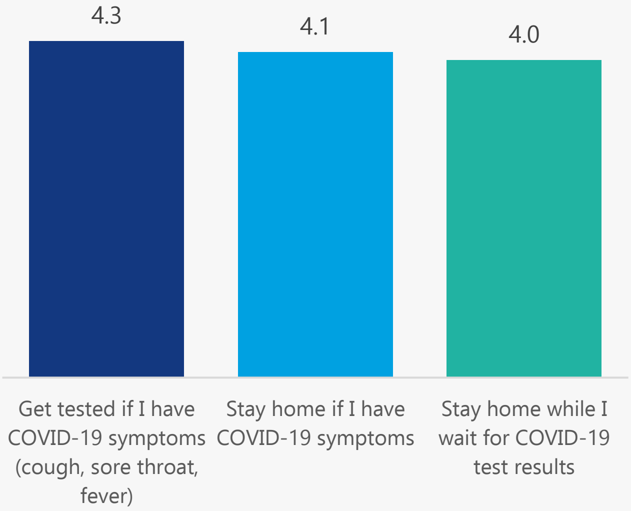


#### Top barriers to getting tested for COVID-19

**50%**

Testing is painful

**50%**

It's too difficult or expensive to get tested

**99%**

I will get tested no matter what

#### Impacts of COVID-19: **Employment**

- **48%** said their employment had changed as a result of COVID-19
- **7%** said they were 'Not at all' or 'A little bit' able to meet their **weekly expenses. 39%** were 'Somewhat' able to, and **54%** said 'Quite a bit' or 'Very much'
- **20%** said they were 'Not at all' or 'A little bit' worried about the **financial problems** they will have in the future as a result of the pandemic. **40%** were 'Somewhat' worried, and **40%** said that they were 'Quite a bit' or 'Very much' worried

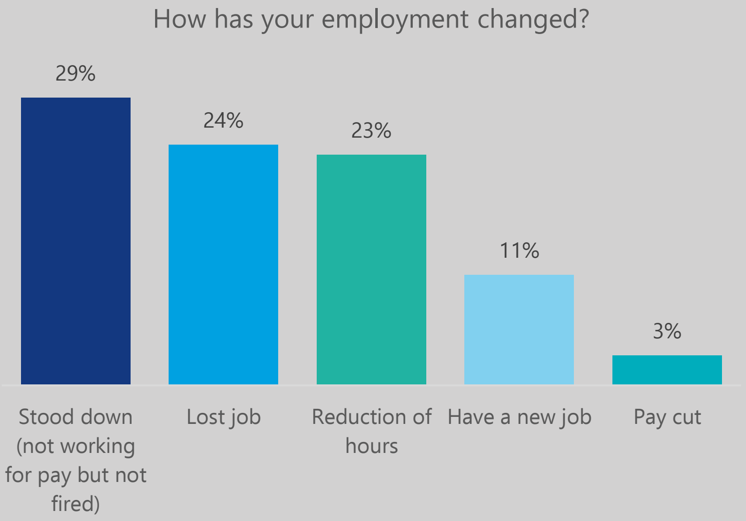


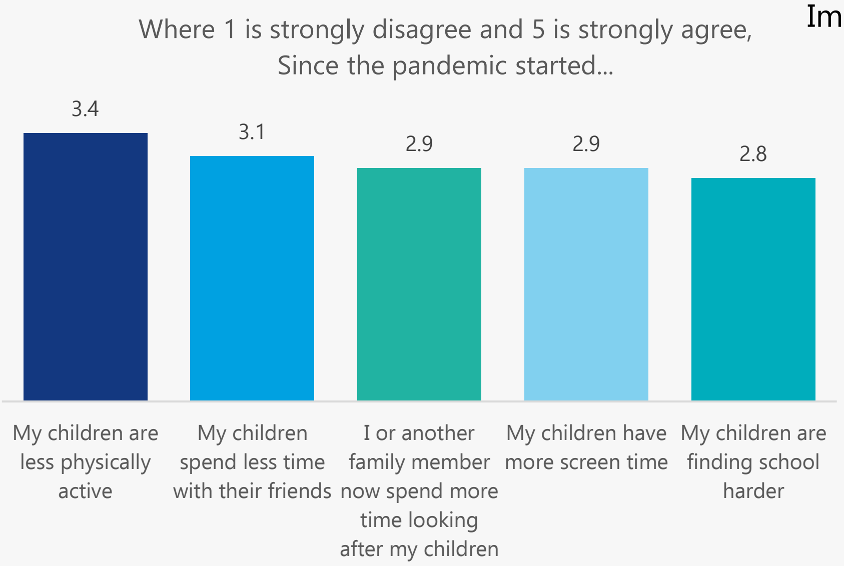


#### Impacts of COVID-19: **Relationships + Children**

- **76%** said COVID-19 has had no effect on their relationship with their partner
- **15%** said COVID-19 has had negative effects on their relationship with their partner
- **9%** said COVID-19 has had positive effects on their relationship with their partner

#### Impacts of COVID-19: **Mental Health**

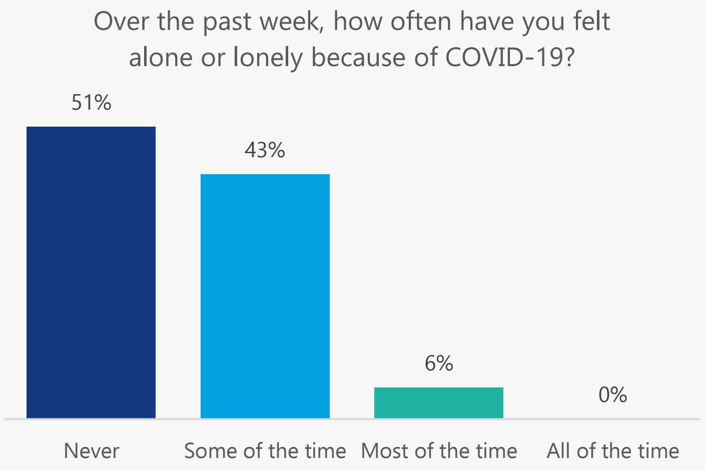


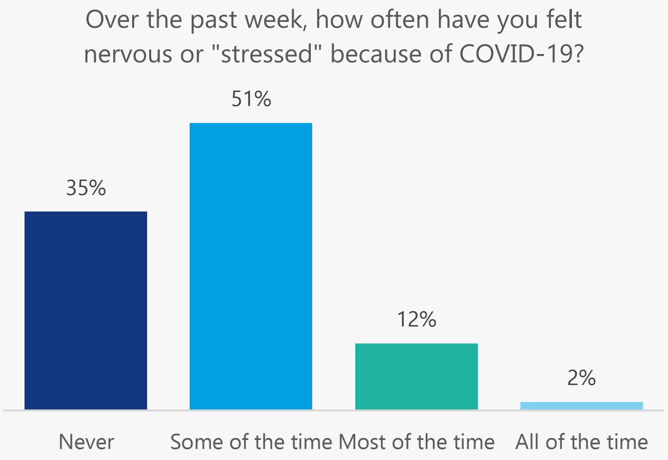


This research was a collaboration between the Sydney Health Literacy Lab (The University of Sydney), Health Literacy Hub (Western Sydney Local Health District), and Western Sydney, South Western Sydney, and Nepean Blue Mountains Local Health Districts. We would like to acknowledge and thank all staff, community members, and participants who made this project possible.

Health Literacy Hub

Sydney Health Literacy Lab

**Notes**

Raw counts are shown in the age and gender figure on page 1. Elsewhere in the report, counts were re-weighted to reflect the population profile in Greater Western Sydney.
This data was collected between 21st March and 9th July 2021. At the time of writing (2nd August 2021), the most recent outbreak commenced on 17th June 2020, and reached 448 cases by day 23 (9th July) when the survey closed.

You can find the University of Sydney Health Literacy Lab at https://sydneyhealthliteracylab.org.au/
The Western Sydney Health Literacy Hub can be found at https://www.healthliteracyhub.org.au/
You can find COVID-19 community resources for Western Sydney LHD at https://www.wslhd.health.nsw.gov.au/
Resources for South Western Sydney LHD at https://www.swslhd.health.nsw.gov.au/
And resources for Nepean Blue Mountains LHD at https://www.nbmlhd.health.nsw.gov.au/

This community summary was prepared by Carys Batcup. To reference the summary, please use this reference:
Ayre J, Muscat DMM, Mac O, Batcup C, Cvejic E, Pickles K, Dolan H, Bonner C, Mouwad D, Zachariah D, Turalic U, Santalucia Y, Chen T, Vasic G, McCaffery K. 2021. COVID-19 Survey Community Summary: Hindi. Available from http://healthliteracyhub.org.au

### COVID-19 Survey

#### Community Summary

### Khmer

A summary of our research into the views of the Khmer speaking community in Greater Western Sydney about COVID-19, conducted between 21/03/21 and 09/07/21

- Average 7.3/10 score for risk perception
- 98% would say Yes to a vaccine
- 89% would get tested 'No matter what'
- Top source for COVID-19 information was official Australian source (92%)

- 63 people who speak Khmer as their main language at home took part
- 56% speak English very well/well (35 out of 63)
- 74% read Khmer very well/well (47 out of 63)
- 66% adequate health literacy (42 out of 63)

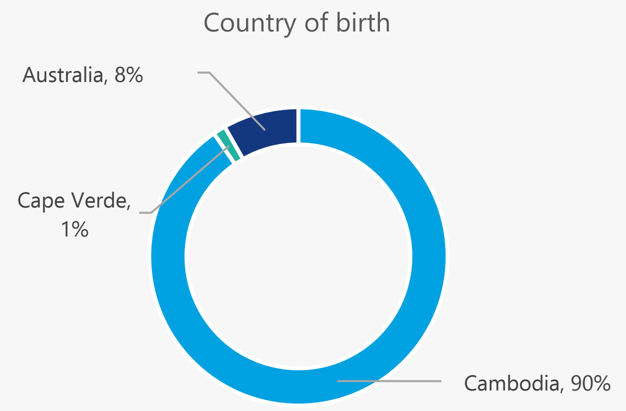


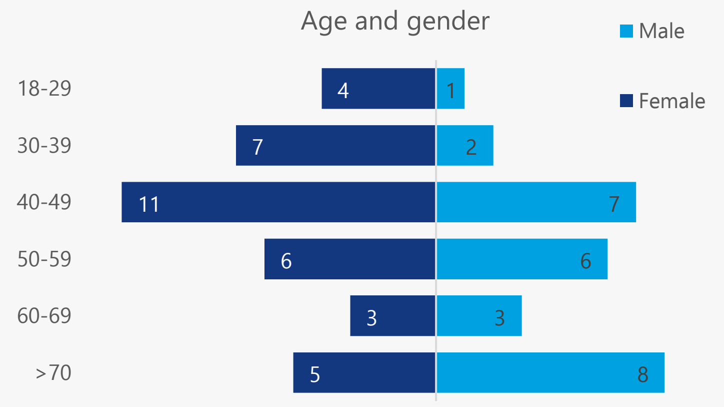


#### Top Sources for COVID-19 Information

1. **Official Australian source (92%):** Australian government websites (76%); Australian public TV (74%); Health professional (52%)
2. **Australian commercial source (82%):** Australian commercial TV (93%); Australian news/magazine website (75%)
3. **Social media (57%):** Facebook (96%); Instagram (74%); YouTube (52%)

**40%** get information about COVID-19 in a language other than English

Average of **2.8** out of 10 for difficulty finding COVID-19 information in Khmer that is easy to understand*

Average of **3.1** out of 10 for difficulty finding information in English that is easy to understand*

**where 10 = extremely difficult*

**98%**

If a COVID-19 vaccine is recommended to me, I will get it

The only barrier given by the one participant who said ‘Not sure’ was ‘I want to wait so that I can see how other countries go first’

- **Risk perception high (average of 7.3 out of 10)**
- **High intentions to perform COVID-19 prevention behaviours (average of 4.9 out of 5)**

On a scale of 0 to 10, how serious a public health problem do you think COVID-19 is currently, in Australia?

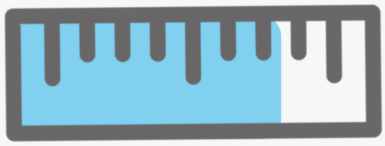

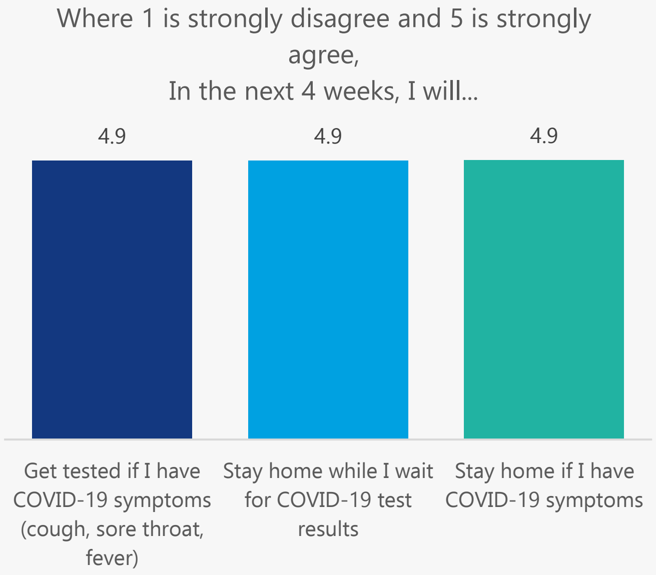


**Barriers to getting tested for COVID-19**

**55%**

I’m worried I will get infected with COVID-19 at the testing clinic

**17%**

Testing is painful

**22%**

I’m worried about what will happen to my visa if I test positive

**6%**

I don’t know how, when and where to get tested

**89%**

I will get tested no matter what

#### Impacts of COVID-19: **Employment**

- **60%** said their employment had changed as a result of COVID-19
- **48%** said they were 'Not at all' or 'A little bit' able to meet their **weekly expenses. 42%** were 'Somewhat' able to, and **10%** said 'Quite a bit' or 'Very much'
- **10%** said they were 'Not at all' or 'A little bit' worried about the **financial problems** they will have in the future as a result of the pandemic. **19%** were 'Somewhat' worried, and **71%** said that they were 'Quite a bit' or 'Very much' worried

How has your employment changed?

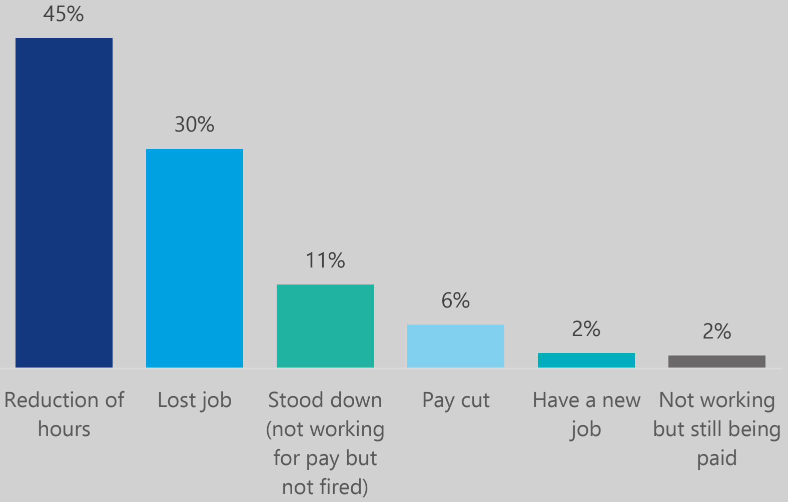


#### Impacts of COVID-19: **Relationships + Children**

- **36%** said COVID-19 has had no effect on their relationship with their partner
- **45%** said COVID-19 has had negative effects on their relationship with their partner
- **19%** said COVID-19 has had some positive effects on their relationship with their partner

Where 1 is strongly disagree and 5 is strongly agree, Since the pandemic started...

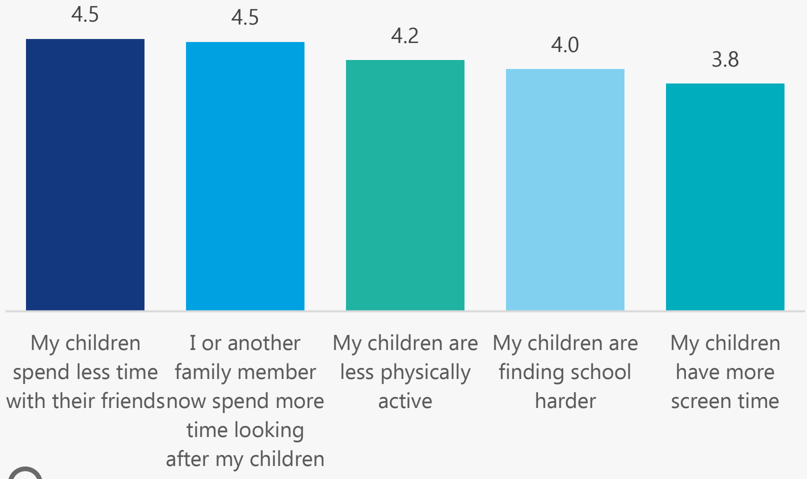


#### Impacts of COVID-19: **Mental Health**

Over the past week, how often have you felt alone or lonely because of COVID-19?

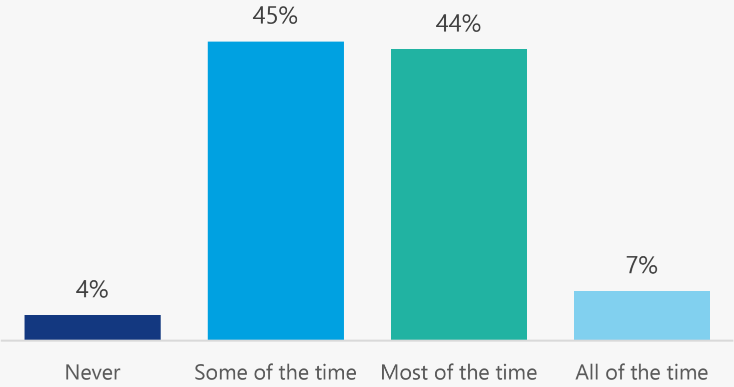


Over the past week, how often have you felt nervous or "stressed" because of COVID-19?

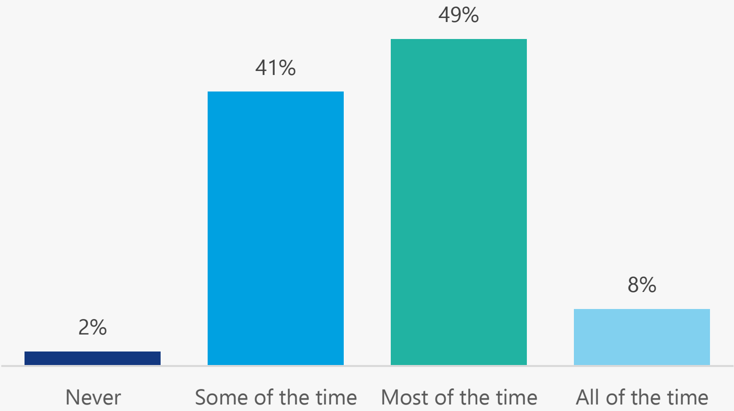


This research was a collaboration between the Sydney Health Literacy Lab (The University of Sydney), Health Literacy Hub (Western Sydney Local Health District), and Western Sydney, South Western Sydney, and Nepean Blue Mountains Local Health Districts. We would like to acknowledge and thank all staff, community members, and participants who made this project possible.

Health Literacy Hub

Sydney Health Literacy Lab

**Notes**

Raw counts are shown in the age and gender figure on page 1. Elsewhere in the report, counts were re-weighted to reflect the population profile in Greater Western Sydney.
This data was collected between 21st March and 9th July 2021. At the time of writing (2nd August 2021), the most recent outbreak commenced on 17th June 2020, and reached 448 cases by day 23 (9th July) when the survey closed.

You can find the University of Sydney Health Literacy Lab at https://sydneyhealthliteracylab.org.au/
The Western Sydney Health Literacy Hub can be found at https://www.healthliteracyhub.org.au/
You can find COVID-19 community resources for Western Sydney LHD at https://www.wslhd.health.nsw.gov.au/
Resources for South Western Sydney LHD at https://www.swslhd.health.nsw.gov.au/
And resources for Nepean Blue Mountains LHD at https://www.nbmlhd.health.nsw.gov.au/

This community summary was prepared by Carys Batcup. To reference the summary, please use this reference:
Ayre J, Muscat DMM, Mac O, Batcup C, Cvejic E, Pickles K, Dolan H, Bonner C, Mouwad D, Zachariah D, Turalic U, Santalucia Y, Chen T, Vasic G, McCaffery K. 2021. COVID-19 Survey Community Summary: Khmer. Available from http://healthliteracyhub.org.au

### COVID-19 Survey

#### Community Summary

### Samoan and Tongan

A summary of our research into the views of the Samoan and Tongan speaking community in Greater Western Sydney about COVID-19, conducted between 21/03/21 and 09/07/21

- Average 4.0/10 score for risk perception
- 29% would say Yes to a vaccine: main barrier is needing more information
- 69% would get tested 'No matter what'
- Top source for COVID-19 information was Australian commercial TV (80%)

- 42 people who speak Samoan or Tongan as their main language at home took part
- 94% speak English very well/well (40 out of 42)
- 92% read Samoan or Tongan very well/well (39 out of 42)
- 98% adequate health literacy (41 out of 42)

Country of birth

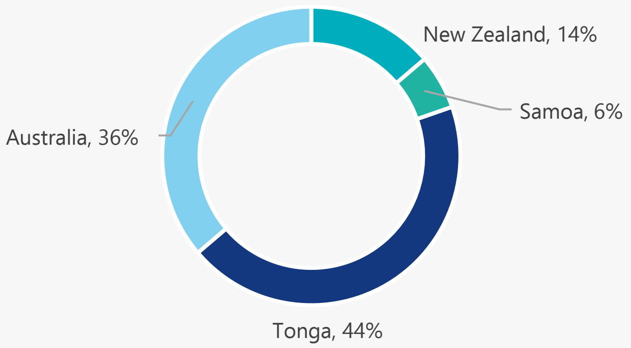


Age and gender

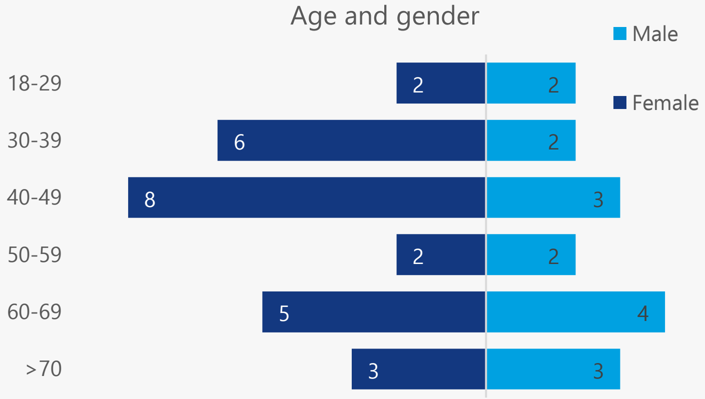


#### Top Sources for COVID-19 Information

1. **Australian commercial source (82%):** Australian commercial TV (98%)
2. **Social media (49%):** Facebook (82%); Instagram (39%); YouTube (23%)
3. **Official Australian source (29%):** Australian government websites (58%); Health professional (53%); Australian public TV (38%)
4. **Community (28%):** Community radio or podcast (73%); Community TV (14%); Religious leader (13%)

**19%** get information about COVID-19 in a language other than English

Average of **6.2** out of 10 for difficulty finding COVID-19 information in Samoan or Tongan that is easy to understand*

Average of **2.2** out of 10 for difficulty finding information in English that is easy to understand*

*where 10 = extremely difficult*

#### Top barriers to getting a COVID-19 vaccine

**29%**

If a COVID-19 vaccine is recommended to me, I will get it

**36%**

I need more information to make a decision

**12%**

I do not think the vaccine will be safe

**14%**

I am worried about side effects

**12%**

I am worried about how the vaccine may affect my other illness

- **Risk perception low (average of 4.0 out of 10)**
- **High intentions to perform COVID-19 prevention behaviours (average of 4.5 out of 5)**

On a scale of 0 to 10, how serious a public health problem do you think COVID-19 is currently, in Australia?

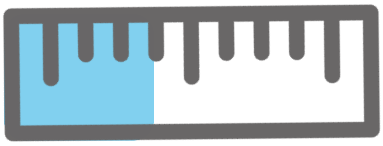

Where 1 is strongly disagree and 5 is strongly agree,
In the next 4 weeks, I will...

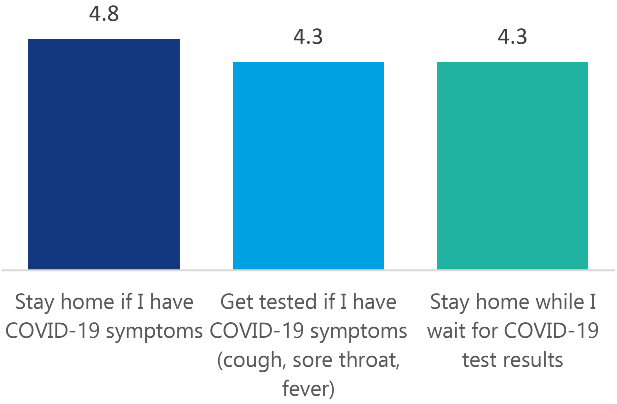


#### Top barriers to getting tested for COVID-19

**53%**

Testing is painful

**15%**

I already had a negative test so I don't need another one

**16%**

I'm worried I will get infected with COVID-19 at the testing clinic

**10%**

I'm worried the results will be on my health record

**69%**

I will get tested no matter what

#### Impacts of COVID-19: **Employment**

- **43%** said their employment had changed as a result of COVID-19
- **37%** said they were 'Not at all' or 'A little bit' able to meet their **weekly expenses. 15%** were 'Somewhat' able to, and **48%** said 'Quite a bit' or 'Very much'
- **35%** said they were 'Not at all' or 'A little bit' worried about the **financial problems** they will have in the future as a result of the pandemic. **9%** were 'Somewhat' worried, and **56%** said that they were 'Quite a bit' or 'Very much' worried

How has your employment changed?

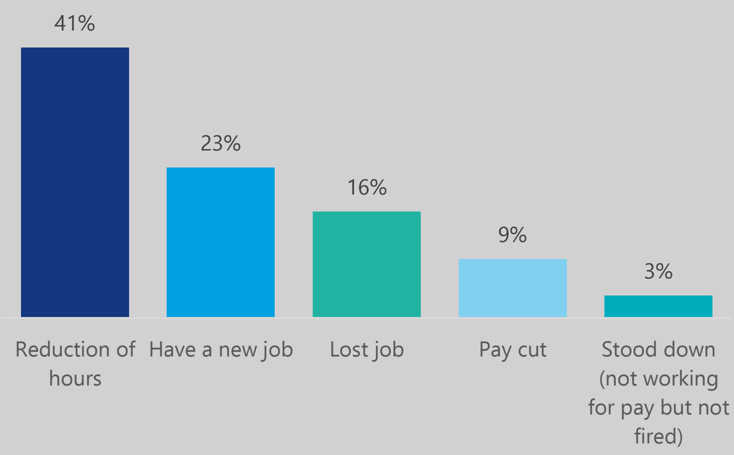


#### Impacts of COVID-19: **Relationships + Children**

Where 1 is strongly disagree and 5 is strongly agree,
Since the pandemic started...

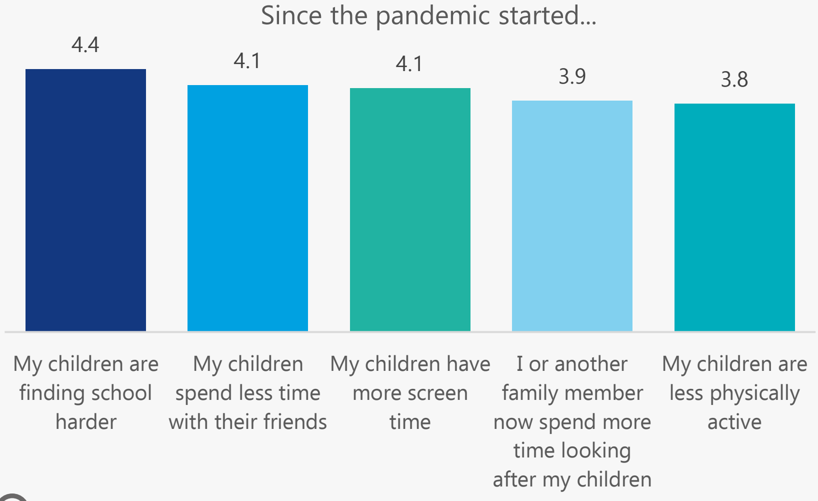


- **32%** said COVID-19 has had no effect on their relationship with their partner
- **68%** said COVID-19 has had negative effects on their relationship with their partner
- **No one** said COVID-19 has had positive effects on their relationship with their partner

#### Impacts of COVID-19: **Mental Health**

Over the past week, how often have you felt alone or lonely because of COVID-19?

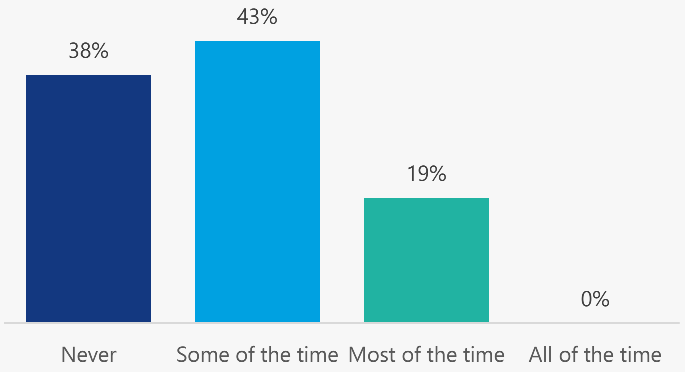


Over the past week, how often have you felt nervous or "stressed" because of COVID-19?

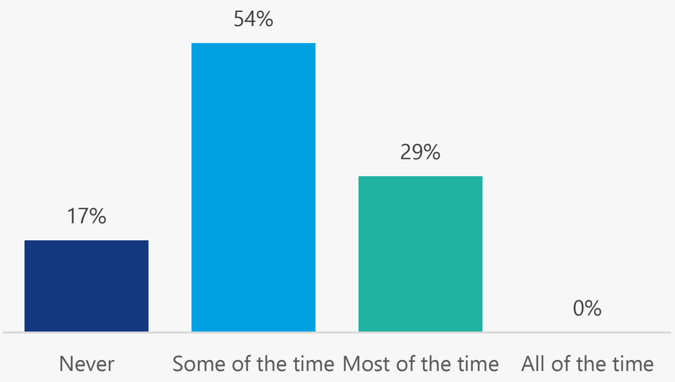


This research was a collaboration between the Sydney Health Literacy Lab (The University of Sydney), Health Literacy Hub (Western Sydney Local Health District), and Western Sydney, South Western Sydney, and Nepean Blue Mountains Local Health Districts. We would like to acknowledge and thank all staff, community members, and participants who made this project possible.

Health Literacy Hub

Sydney Health Literacy Lab

**Notes**

Raw counts are shown in the age and gender figure on page 1. Elsewhere in the report, counts were re-weighted to reflect the population profile in Greater Western Sydney.

This data was collected between 21st March and 9th July 2021. At the time of writing (2nd August 2021), the most recent outbreak commenced on 17th June 2020, and reached 448 cases by day 23 (9th July) when the survey closed.

You can find the University of Sydney Health Literacy Lab at https://sydneyhealthliteracylab.org.au/

The Western Sydney Health Literacy Hub can be found at https://www.healthliteracyhub.org.au/

You can find COVID-19 community resources for Western Sydney LHD at https://www.wslhd.health.nsw.gov.au/

Resources for South Western Sydney LHD at https://www.swslhd.health.nsw.gov.au/

And resources for Nepean Blue Mountains LHD at https://www.nbmlhd.health.nsw.gov.au/

This community summary was prepared by Carys Batcup. To reference the summary, please use this reference:

Ayre J, Muscat DMM, Mac O, Batcup C, Cvejic E, Pickles K, Dolan H, Bonner C, Mouwad D, Zachariah D, Turalic U, Santalucia Y, Chen T, Vasic G, McCaffery K. 2021. COVID-19 Survey Community Summary: Samoan and Tongan. Available from http://healthliteracyhub.org.au
